# Temporal Trends in Racial and Ethnic Disparities in Multimorbidity Prevalence in the United States, 1999–2018

**DOI:** 10.1101/2021.08.25.21262641

**Authors:** César Caraballo, Shiwani Mahajan, Daisy Massey, Yuan Lu, Chima D. Ndumele, Elizabeth E. Drye, Jeph Herrin, Harlan M. Krumholz

## Abstract

**Background:** Disparities in multimorbidity prevalence indicate health inequalities. This study sought to determine if gaps in multimorbidity prevalence by race and ethnicity are decreasing over time.

**Methods and findings:** Serial cross-sectional analysis of the National Health Interview Survey from years 1999 to 2018. The study included individuals ≥18 years old, categorized by self-reported race, ethnicity, age, and income. The main outcomes were temporal trends in multimorbidity prevalence based on the self-reported presence of ≥2 of 9 chronic conditions. The study sample included 596,355 individuals (4.7% Asian, 11.8% Black, 13.8% Latino/Hispanic, and 69.7% White). In 1999, the estimated prevalence of multimorbidity was 5.9% among Asian, 17.4% among Black, 10.7% among Latino/Hispanic, and 13.5% among White individuals. Prevalence increased for all racial/ethnic groups during the study period (P<0.005 for each), with no significant change in the differences between them. In 2018, when compared with White people, multimorbidity was more prevalent among Black individuals (+2.5 percent points [95% CI: +0.5, +4.6]) and less prevalent among Asian and Latino individuals (−6.6 points [-8.8, -4.4] and -2.1 points [-4.0, -0.2], respectively). The estimated prevalence of reporting at least 1, 2, 3, or 4 conditions all increased with age across all groups, with Black individuals having the greatest prevalence in each age category. Among those aged ≥30 years, Black people had multimorbidity rates equivalent to those of Latino/Hispanic and White people aged 5 years older, and Asian people aged 10 years older. Notably, the prevalence of multimorbidity diverged between Black people and White people among those aged 30 years, reached its maximum difference of 10.1 percentage points (95% CI: 8.2, 12.1) among those aged 60, and converged to reach similar rates among those aged ≥80.

**Conclusions:** From 1999 to 2018, a period of increasing multimorbidity prevalence for all the groups studied, there has not been any significant progress in eliminating disparities between Black and White people. The disparity began among young adults. Public health interventions that prevent the onset of chronic conditions in early life may be needed to eliminate these disparities.

## INTRODUCTION

In 2010, the US Department of Health and Human Services published a public health framework for managing patients with multimorbidity, defined as the presence of 2 or more concurrent chronic conditions in an individual, highlighting the importance of identifying and addressing disparities in its prevalence.[1, 2] Despite this report, differences in the prevalence of multimorbidity remain understudied. Recent studies indicate that multimorbidity is more prevalent among Black people compared with White people and that its overall prevalence is increasing in the US.[3-5] Given the lack of a comprehensive investigation of trends in multimorbidity over the past 2 decades, especially by race and ethnicity, it is unknown if the US has made progress in narrowing the gaps. The examination of trends in multimorbidity has implications about accountability for what the health system in the US has achieved in population health and health equity over the past 20 years, and what it will face in the future.

Accordingly, we describe the 20-year trends in the prevalence of multimorbidity by race and ethnicity, estimating the annual differences between groups. We also evaluate whether the effect of race and ethnicity on multimorbidity prevalence varied by age, describing the age groups among which these differences begin and are the highest. Finally, considering that income is one of the most critical determinants of health, we stratified the main analyses by income level, which can provide insight into the role of income inequality in the disparities studied.

## METHODS

### Data Source

We used data from the annual National Health Insurance Survey (NHIS) from 1999 to 2018. The NHIS is a cross-sectional representative survey of the non-institutionalized population in the US that describes the health conditions, health insurance coverage and utilization, and health behaviors of recipients. The NHIS uses a complex multistage area probability design that accounts for non-response and allows for nationally representative estimates, including for underrepresented groups.[6] We obtained the data from the Integrated Public Use Microdata Series Health Surveys (https://nhis.ipums.org/).[7] The Institutional Review Board at Yale University exempted the study from review. Author César Caraballo had full access to the data and takes responsibility for the accuracy of the analysis. The code used in the analysis is publicly available at http://doi.org/10.3886/E151983V1.

### Study Population

We included individuals ≥18 years old from years 1999 to 2018 of the Sample Adult Core file, which contains responses from an in-depth questionnaire administered to a randomly selected adult from each family. We excluded responses with missing information about chronic conditions. Due to small numbers, we also excluded those who did not identify as non-Hispanic Asian, non-Hispanic Black/African American, Latino/Hispanic, or non-Hispanic White (details in Results section). The mean conditional and final response rate of the Sample Adult Core survey during the study period was 81% and 64.8%, respectively.

#### Demographic Variables

Race was determined by the participants’ self-reported primary race selection. Latino/Hispanic ethnicity was defined as answering “Yes” to the question, “Do you consider yourself Latino/Hispanic?” Participants were divided into 4 mutually exclusive subgroups: non-Hispanic Asian (Asian), non-Hispanic Black/African American (Black), Latino/Hispanic, and non-Hispanic White (White).

Respondent characteristics included in the analyses were self-reported age, sex, geographic region (Northeast, North Central/Midwest, South, West), and income. Age was also categorized by 5-year groups for stratification and reporting. Consistent with prior NHIS studies, family income was categorized according to its relative percentage of the federal poverty level from the US Census Bureau[8] into middle/high income (≥200% of the federal poverty level) and low income (<200%).[9-11] Other sociodemographic and clinical variables were used to describe the characteristics of the population.

### Chronic Conditions

We included 9 chronic conditions that the US Centers for Disease Control and Prevention consistently included in the NHIS across the entire study period. These conditions were among those listed by the US Department of Health and Human Services Office of the Assistant Secretary of Health conceptual framework for studying multiple chronic conditions. We identified individuals with these conditions if they had ever been told by a health care professional that they had diabetes, hypertension, asthma, stroke, cancer, chronic obstructive pulmonary disease (emphysema or chronic bronchitis), or heart disease (coronary artery disease, myocardial infarction, angina, or other heart conditions), or had been told in the past 12 months that they had weak/failing kidneys or any liver condition. Participants were then classified as having 0, 1, 2, 3, or ≥4 chronic conditions. We defined multimorbidity as the presence of ≥2 of the 9 chronic conditions.[1, 12-14]

### Statistical Analysis

We summarized the general characteristics of respondents by racial/ethnic group. Then, we estimated the annual multimorbidity prevalence for each group using multivariable logistic regression models adjusting for age, sex, and region (details in Supporting Information Methods). To assess the trends in the prevalence of each of the 9 conditions, we used the same approach as for multimorbidity overall.

We subtracted the annual prevalence among White individuals from that year’s prevalence among Asian, Black, and Latino/Hispanic individuals, also constructing SE for the differences, to quantify the racial and ethnic gap. Using these prevalences and gaps, we estimated trends over the study period by fitting weighted linear regression models (details in Supporting Information Methods). We also used a z-test to estimate the absolute difference between 1999 and 2018 in the multimorbidity prevalence within each race and ethnic group, and the differences between groups.

Then, we estimated the adjusted annual mean number of chronic conditions by race and ethnicity using linear regressions. We also used ordered logistic regression models to estimate the proportion of individuals with 0, 1, 2, 3, or ≥4 chronic conditions over the years. In these models (linear and ordered logit), the number of conditions was the dependent variable and age, sex, region, and an indicator for each year were the independent variables, obtaining the estimates for each race/ethnicity group separately.

To assess if the association between race and ethnicity and multimorbidity differs by income, we replicated the main analysis by income group. Due to high rates of missing income information from non-response, the publicly available NHIS data included multiply imputed income variables for respondents who did not report income. Thus, we followed the recommendations from the National Center for Health Statistics for multiply imputed data analysis (details in Supporting Information Methods).[15]

To evaluate if the association between race and ethnicity and multimorbidity differs by age, we used a logistic regression model with multimorbidity as the dependent variable, region, sex, age and race/ethnicity as independent variables, and an interaction term between age and race and ethnicity. We reported the interaction term odds ratio (OR) for each race and ethnicity group. We replicated this using an ordered logit model and the number of chronic conditions as the dependent variable, and age group, region, and sex as the independent variables to estimate the percentage with 0, 1, 2, 3, and ≥4 conditions in each age group by race and ethnicity. We reported the findings from these analyses based on the approach of Barnett et al.[16]

Lastly, we performed a sensitivity analysis including arthritis in the list of conditions using data from 2002 to 2018, the period in which the condition was available and ascertained consistently. The purpose of this sensitivity analysis was to compare our multimorbidity prevalence estimates with those obtained from NHIS studies that included arthritis in their calculation.[17-19]

For all analyses, a 2-sided P-value <0.05 was used to determine statistical significance. All analyses were performed using Stata SE version 17.0 (StataCorp, College Station, TX). All results are reported with 95% CIs. All analyses incorporated strata and weights to produce nationally representative estimates using the Stata -*svy-* command for structured survey data. All person-weights were pooled and divided by the number of years studied, following guidance from the NHIS.[20]

## RESULTS

### Population characteristics

Of a total of 603,140 adults interviewed, we excluded 112 with missing race and ethnicity information and 119 with missing chronic conditions information. Due to small representation, we also excluded 6673 with ethnicity other than Latino/Hispanic who also did not select a primary race (n=2114) or who selected primary race as Alaskan Native or American Indian (n=4355) or Other (n=204) (S1 Fig). The final study sample comprised 596,236 adults with an estimated mean age of 46.2 [SE, 0.07] years of which 51.8% [95% CI: 51.7, 52.0] were women. Of these, 4.7% (95% CI: 4.6%, 4.8%) identified as Asian, 11.8% (95% CI: 11.5%, 12.1%) as Black, 13.8% (95% CI: 13.5%, 14.2%) as Latino/Hispanic, and 69.7% (95% CI: 69.3%, 70.2%) as White. The overall estimated prevalence of multimorbidity was 19.9% (95% CI: 19.7%, 20.1%), increasing from 15.2% (95% CI: 14.8%, 15.7%) in 1999 to 22.7% (95% CI: 22.1%, 23.4%) in 2018. The general characteristics of the study population are shown in Table 1, S1 Table, and S2 Fig. The overall unadjusted distribution of number of chronic conditions by race and ethnicity is shown in S3 Fig, and the annual adjusted prevalence of each of the conditions by race and ethnicity is shown in S4 Fig.

**Table 1.** General Characteristics of the Study Population by Race and Ethnicity.

|  | Asian |  |  | Black |  |  | Latino/Hispanic |  |  | White |  |  |
| --- | --- | --- | --- | --- | --- | --- | --- | --- | --- | --- | --- | --- |
|  | 1999–2000 | 2008–2009 | 2017–2018 | 1999–2000 | 2008–2009 | 2017–2018 | 1999–2000 | 2008–2009 | 2017–2018 | 1999–2000 | 2008–2009 | 2017–2018 |
| Sample size, n<br>[Total=596,236] | n=1597 | n=2828 | n=2640 | n=8758 | n=7804 | n=5881 | n=10,305 | n=8838 | n=6422 | n=41,872 | n=29,553 | n=36,401 |
| **Age in years** | 39 (28–55) | 41 (30–55) | 43 (32–57) | 40 (29–52) | 42 (29–55) | 43 (30–58) | 37 (27–49) | 37 (28–50) | 39 (28–53) | 44 (33–59) | 47 (33–61) | 50 (34–64) |
| **Age category** |  |  |  |  |  |  |  |  |  |  |  |  |
| 18–39 years | 52.4 (49.2, 55.6) | 45.9 (43.4, 48.5) | 41.9 (39.3, 44.6) | 49.8 (48.3, 51.3) | 45.1 (43.4, 46.8) | 43.7 (41.8, 45.6) | 56.9 (55.4, 58.3) | 54.6 (53.0, 56.1) | 50.1 (48.4, 51.9) | 39.5 (38.9, 40.2) | 34.9 (34.0, 35.8) | 33.3 (32.6, 34.1) |
| 40–64 years | 38.2 (35.1, 41.4) | 40.8 (38.2, 43.5) | 42.5 (40.2, 44.9) | 38.5 (37.1, 39.8) | 42.9 (41.4, 44.4) | 41.1 (39.3, 42.9) | 34.1 (33.0, 35.3) | 37.1 (35.7, 38.5) | 39.1 (37.6, 40.6) | 42.2 (41.5, 42.8) | 45.8 (45.0, 46.5) | 42.3 (41.6, 43.0) |
| ≥65 years | 9.4 (7.8, 11.4) | 13.3 (11.8, 14.9) | 15.5 (13.9, 17.4) | 11.8 (11.0, 12.6) | 12.0 (11.2, 12.9) | 15.3 (14.2, 16.3) | 9.0 (8.1, 10.0) | 8.4 (7.7, 9.1) | 10.8 (9.9, 11.8) | 18.3 (17.8, 18.8) | 19.3 (18.7, 20.0) | 24.4 (23.7, 25.1) |
| **Women** | 51.4 (48.1, 54.6) | 52.6 (50.3, 54.9) | 53.3 (51.1, 55.6) | 55.6 (54.1, 57.0) | 55.4 (53.9, 56.9) | 54.5 (52.8, 56.3) | 50.6 (49.4, 51.8) | 48.6 (47.2, 50.0) | 50.3 (48.7, 51.9) | 51.9 (51.3, 52.4) | 51.6 (51.0, 52.3) | 51.4 (50.7, 52.0) |
| **US Citizenship**<br>[n=594,859] | 57.9 (54.3, 61.4) | 69.6 (66.8, 72.2) | 70.5 (67.9, 73.1) | 95.4 (94.7, 96.0) | 95.0 (94.2, 95.6) | 94.9 (93.8, 95.9) | 62.6 (60.5, 64.7) | 62.2 (60.2, 64.1) | 72.3 (70.6, 74.0) | 98.4 (98.2, 98.5) | 98.5 (98.3, 98.7) | 98.5 (98.3, 98.7) |
| **Education level**<br>[n=591,667] |  |  |  |  |  |  |  |  |  |  |  |  |
| Less than high school | 11.4 (9.4, 13.7) | 9.2 (7.9, 10.7) | 7.9 (6.6, 9.5) | 24.4 (23.1, 25.8) | 17.7 (16.4, 19.1) | 14.2 (12.9, 15.7) | 44.7 (43.0, 46.3) | 38.5 (37.0, 40.1) | 27.8 (26.0, 29.7) | 13.4 (12.9, 13.9) | 10.5 (10.0, 11.1) | 7.4 (6.9, 7.9) |
| High school diploma /GED | 18.1 (16.1, 20.2) | 16.3 (14.4, 18.3) | 15.3 (13.5, 17.4) | 31.5 (30.2, 32.8) | 30.6 (29.2, 32.1) | 28.9 (27.3, 30.5) | 24.3 (23.3, 25.4) | 26.6 (25.2, 28.0) | 26.7 (25.1, 28.3) | 31.7 (31.0, 32.4) | 28.2 (27.5, 28.9) | 23.6 (22.9, 24.4) |
| Some college | 24.2 (21.7, 26.9) | 24.4 (22.2, 26.7) | 20.8 (18.8, 22.9) | 29.7 (28.3, 31.0) | 34.0 (32.5, 35.5) | 32.9 (31.2, 34.7) | 21.7 (20.5, 23.0) | 22.6 (21.4, 23.8) | 28.1 (26.5, 29.7) | 29.5 (29.0, 30.0) | 31.5 (30.9, 32.2) | 31.4 (30.6, 32.1) |
| ≥Bachelor’s degree | 46.4 (43.3, 49.5) | 50.1 (46.8, 53.5) | 56.0 (52.9, 59.0) | 14.5 (13.4, 15.7) | 17.7 (16.5, 18.9) | 24.1 (22.2, 26.0) | 9.4 (8.5, 10.3) | 12.4 (11.4, 13.4) | 17.5 (16.0, 19.0) | 25.4 (24.7, 26.1) | 29.8 (28.9, 30.7) | 37.6 (36.5, 38.7) |
| **Income <200% of Federal Poverty Limit<sup>a</sup>** | 28.0 (23.9, 32.5) | 27.1 (23.9, 30.5) | 26.2 (22.9, 29.9) | 45.6 (43.3, 47.8) | 45.0 (42.7, 47.2) | 44.4 (41.4, 47.4) | 51.0 (49.0, 53.1) | 50.3 (48.1, 52.5) | 46.7 (44.1, 49.3) | 23.0 (22.2, 23.8) | 23.9 (22.8, 25.1) | 21.3 (20.3, 22.3) |
| **Uninsured at the time of interview**<br>[n=594,011] | 17.4 (15.2, 19.8) | 13.9 (12.2, 15.8) | 6.3 (5.3, 7.6) | 20.3 (19.2, 21.4) | 20.8 (19.6, 22.0) | 12.1 (10.8, 13.5) | 36.3 (34.5, 38.1) | 38.9 (37.0, 40.8) | 24.0 (22.2, 26.0) | 11.0 (10.6, 11.4) | 12.3 (11.8, 12.9) | 6.5 (6.2, 6.9) |
| **US region of residence<sup>b</sup>** |  |  |  |  |  |  |  |  |  |  |  |  |
| Northeast | 21.2 (18.7, 24.0) | 18.8 (16.4, 21.5) | 20.8 (16.9, 25.4) | 18.0 (16.7, 19.4) | 16.4 (14.8, 18.2) | 16.0 (13.5, 18.8) | 15.7 (14.5, 17.1) | 13.4 (11.7, 15.2) | 13.5 (11.2, 16.2) | 20.2 (19.5, 21.0) | 18.3 (17.3, 19.3) | 19.2 (17.5, 21.1) |
|  | 1999–2000 | 2008–2009 | 2017–2018 | 1999–2000 | 2008–2009 | 2017–2018 | 1999–2000 | 2008–2009 | 2017–2018 | 1999–2000 | 2008–2009 | 2017–2018 |
| Midwest | 14.7 (11.9, 18.0) | 15.0 (12.7, 17.7) | 11.8 (9.5, 14.6) | 18.8 (17.2, 20.5) | 19.3 (17.3, 21.5) | 15.2 (13.0, 17.8) | 7.9 (6.9, 9.1) | 9.5 (8.1, 11.2) | 9.4 (7.6, 11.6) | 29.4 (28.5, 30.3) | 28.6 (27.2, 30.0) | 27.4 (25.7, 29.3) |
| South | 18.9 (16.4, 21.8) | 19.3 (17.2, 21.7) | 25.5 (21.6, 30.0) | 55.7 (53.4, 57.9) | 56.1 (53.5, 58.7) | 60.5 (56.7, 64.3) | 35.4 (33.2, 37.8) | 35.4 (33.4, 37.4) | 37.1 (32.5, 42.0) | 33.9 (33.0, 34.8) | 33.8 (32.3, 35.4) | 33.0 (30.9, 35.2) |
| West | 45.2 (41.2, 49.2) | 46.8 (43.6, 50.1) | 41.8 (36.7, 47.1) | 7.6 (6.7, 8.5) | 8.1 (7.3, 9.1) | 8.3 (6.9, 9.9) | 40.9 (38.6, 43.3) | 41.7 (39.4, 44.1) | 40.0 (35.3, 44.8) | 16.5 (15.8, 17.2) | 19.3 (18.3, 20.4) | 20.4 (18.3, 22.7) |
| **Married or living with partner**<br>[n=593,807] | 64.4 (61.3, 67.3) | 64.1 (61.2, 66.8) | 65.0 (62.5, 67.5) | 37.4 (35.9, 38.9) | 35.2 (33.6, 36.8) | 32.6 (31.0, 34.3) | 58.5 (57.4, 59.6) | 54.4 (52.8, 56.0) | 49.3 (47.6, 50.9) | 61.6 (60.8, 62.3) | 57.9 (56.9, 58.8) | 56.4 (55.6, 57.1) |
| **Employment status**<br>[n=595,478] |  |  |  |  |  |  |  |  |  |  |  |  |
| With a job/Working | 66.1 (63.4, 68.6) | 65.1 (62.6, 67.5) | 67.2 (64.9, 69.3) | 64.1 (62.6, 65.5) | 60.7 (59.2, 62.3) | 61.1 (59.0, 63.1) | 66.3 (65.0, 67.5) | 64.2 (62.9, 65.5) | 66.5 (64.7, 68.3) | 65.7 (65.0, 66.4) | 62.5 (61.6, 63.4) | 62.1 (61.3, 63.0) |
| Not in labor force | 31.8 (29.2, 34.5) | 30.4 (28.0, 32.8) | 30.0 (27.8, 32.3) | 32.0 (30.6, 33.5) | 31.3 (29.9, 32.8) | 32.4 (30.5, 34.4) | 31.2 (30.0, 32.4) | 28.9 (27.8, 30.1) | 29.9 (28.0, 31.8) | 32.9 (32.3, 33.6) | 33.4 (32.5, 34.3) | 35.4 (34.6, 36.2) |
| Unemployed | 2.2 (1.5, 3.1) | 4.6 (3.7, 5.6) | 2.8 (2.1, 3.8) | 3.9 (3.4, 4.6) | 7.9 (7.2, 8.8) | 6.5 (5.7, 7.4) | 2.5 (2.2, 2.9) | 6.9 (6.2, 7.7) | 3.6 (3.0, 4.3) | 1.4 (1.3, 1.5) | 4.1 (3.8, 4.4) | 2.5 (2.3, 2.7) |
| **Chronic conditions** |  |  |  |  |  |  |  |  |  |  |  |  |
| Asthma | 5.5 (4.4, 6.9) | 9.1 (7.8, 10.6) | 8.4 (7.2, 9.9) | 9.0 (8.2, 9.9) | 13.9 (12.9, 13.8) | 15.1 (14.0, 16.2) | 7.1 (6.5, 7.7) | 10.0 (9.2, 10.8) | 11.6 (10.5, 12.8) | 9.2 (8.9, 9.5) | 13.3 (12.9, 13.8) | 13.9 (13.5, 14.3) |
| Cancer | 1.7 (1.1, 2.7) | 3.0 (2.4, 3.7) | 4.3 (3.4, 5.3) | 2.9 (2.5, 3.4) | 3.8 (3.4, 4.4) | 4.8 (4.3, 5.4) | 2.2 (1.9, 2.5) | 2.7 (2.3, 3.2) | 3.4 (2.9, 3.8) | 7.8 (7.5, 8.1) | 10.2 (9.8, 10.6) | 12.3 (11.9, 12.8) |
| COPD | 1.6 (1.1, 2.4) | 1.8 (1.3, 2.6) | 2.0 (1.4, 2.7) | 4.3 (3.8, 4.7) | 4.5 (4.0, 5.1) | 4.4 (3.7, 5.1) | 2.9 (2.5, 3.4) | 2.8 (2.4, 3.4) | 2.8 (2.4, 3.3) | 6.2 (5.9, 6.5) | 6.5 (6.2, 6.9) | 5.3 (5.0, 5.6) |
| Diabetes | 3.9 (3.0, 5.1) | 7.4 (6.2, 8.8) | 8.9 (7.7, 10.3) | 8.2 (7.5, 8.9) | 11.3 (10.4, 12.2) | 11.7 (10.8, 12.7) | 6.3 (5.8, 6.9) | 8.7 (8.0, 9.6) | 10.5 (9.6, 11.4) | 5.2 (5.0, 5.4) | 8.2 (7.9, 8.6) | 9.1 (8.7, 9.4) |
| Heart disease | 4.6 (3.6, 5.9) | 4.9 (4.0, 6.2) | 6.6 (5.6, 7.7) | 8.9 (8.2, 9.7) | 9.9 (9.1, 10.8) | 9.8 (9.0, 10.7) | 6.0 (5.4, 6.6) | 6.1 (5.5, 6.7) | 6.4 (5.7, 7.2) | 12.1 (11.7, 12.4) | 13.8 (13.2, 14.3) | 14.0 (13.6, 14.5) |
| Hypertension | 14.9 (12.9, 17.2) | 22.3 (20.2, 24.6) | 24.2 (22.2, 26.3) | 28.8 (27.6, 30.1) | 35.4 (34.0, 36.9) | 37.6 (35.9, 39.3) | 15.3 (14.4, 16.2) | 20.0 (18.9, 21.1) | 22.3 (21.0, 23.8) | 23.0 (22.5, 23.4) | 30.4 (29.7, 31.1) | 32.7 (31.9, 33.4) |
| Kidney disease | 0.8 (0.5, 1.5) | 1.1 (0.8, 1.7) | 1.8 (1.2, 2.6) | 2.0 (1.7, 2.5) | 2.2 (1.7, 2.7) | 2.6 (2.2, 3.1) | 1.6 (1.3, 1.9) | 1.8 (1.6, 2.1) | 2.0 (1.6, 2.5) | 1.3 (1.2, 1.5) | 1.8 (1.6, 2.0) | 2.3 (2.1, 2.5) |
| Liver disease | 1.1 (0.5, 2.3) | 1.3 (0.9, 2.0) | 1.7 (1.2, 2.4) | 1.0 (0.8, 1.3) | 1.0 (0.7, 1.3) | 1.2 (0.9, 1.5) | 1.0 (0.8, 1.2) | 1.8 (1.5, 2.1) | 2.4 (2.0, 2.9) | 1.0 (0.9, 1.1) | 1.5 (1.3, 1.7) | 1.8 (1.6, 2.0) |
| Stroke | 0.8 (0.6, 1.2) | 1.2 (0.8, 1.7) | 1.9 (1.4, 2.5) | 2.8 (2.4, 3.2) | 3.3 (2.8, 3.8) | 4.0 (3.5, 4.6) | 1.1 (0.9, 1.4) | 1.6 (1.3, 1.9) | 2.1 (1.7, 2.6) | 2.2 (2.1, 2.3) | 3.0 (2.8, 3.2) | 3.3 (3.1, 3.6) |
| **Current smoker** | 14.9 (12.9, 17.2) | 11.0 (9.5, 12.7) | 7.2 (6.0, 8.5) | 23.8 (22.6, 25.1) | 21.3 (20.0, 22.7) | 14.7 (13.5, 16.1) | 18.3 (17.4, 19.3) | 15.2 (14.1, 16.2) | 9.8 (9.0, 10.8) | 24.2 (23.7, 24.8) | 22.1 (21.4, 22.8) | 15.1 (14.6, 15.7) |
|  | 1999–2000 | 2008–2009 | 2017–2018 | 1999–2000 | 2008–2009 | 2017–2018 | 1999–2000 | 2008–2009 | 2017–2018 | 1999–2000 | 2008–2009 | 2017–2018 |
| **Flu vaccine in past 12 months** | 26.1 (23.3, 29.2) | 33.4 (31.2, 35.7) | 48.2 (45.6, 50.7) | 20.6 (19.7, 21.5) | 26.3 (25.0, 27.7) | 35.2 (33.4, 37.1) | 18.6 (17.5, 19.6) | 22.1 (20.9, 23.2) | 34.9 (33.3, 36.5) | 30.7 (30.0, 31.3) | 36.9 (36.2, 37.6) | 47.1 (46.4, 47.9) |
| **Obese (BMI $\geq 30$ kg/m<sup>2</sup>)** | 6.1 (4.9, 7.7) | 8.7 (7.1, 10.5) | 12.0 (10.5, 13.6) | 29.1 (28.0, 30.2) | 37.1 (35.8, 38.4) | 39.4 (37.8, 41.1) | 23.0 (21.9, 24.1) | 31.2 (29.9, 32.4) | 34.0 (32.5, 35.6) | 20.2 (19.7, 20.6) | 26.1 (25.4, 26.8) | 30.3 (29.6, 31.0) |
| Data are presented as % (95% CI) for categorical variables and median (P25–P75) for continuous variables. All percentages are unadjusted and weighted. |  |  |  |  |  |  |  |  |  |  |  |  |
| <sup>a</sup> Annual family income was categorized relative to the respective year’s Federal Poverty Level from the US Census Bureau into middle/high income ($\geq 200\%$) and low income ($<200\%$). The weighted proportion of individuals with annual income $<200\%$ of the Federal Poverty Limit was estimated using multiple imputation.<br><sup>b</sup> Based on the US Census Bureau recognized region where the housing unit of the survey participant was located. | | | | | | | | | | | | |
| Abbreviations: BMI, body mass index; CI, confidence interval; COPD, chronic obstructive pulmonary disease; GED, general equivalency diploma. |  |  |  |  |  |  |  |  |  |  |  |  |

### Temporal Multimorbidity Trends, 1999 to 2018

In 1999, the estimated age-, sex-, and region-adjusted prevalence of multimorbidity was 5.9% (95% CI: 4.3, 8.0) among Asian people, 17.2% (95% CI: 17.7, 18.8) among Black people, 10.5% (95% CI: 9.3, 11.7) among Latino/Hispanic people, and 13.2% (95% CI: 12.8, 13.7) among White people (Fig 1). Between 1999 and 2018, the adjusted multimorbidity prevalence increased significantly within each race and ethnicity group (P=0.001 for Black people and P<0.001 for Asian, Latino/Hispanic, and White people). There were no significant changes in the difference between groups (Table 2). In 2018, when compared with the adjusted multimorbidity prevalence among White people (18.7% [95% CI: 18.0, 19.5]), the adjusted multimorbidity prevalence among Black people was estimated to be 2.5 percentage points higher (95% CI: 0.5, 4.6; P=0.02), whereas it was lower among Asian and Latino/Hispanic people by 6.6 points (95% CI: -8.8, -4.4; P<0.001) and 2.1 points (95% CI: -4.0, -0.2; P=0.03), respectively. Throughout the study period, the adjusted prevalence of multimorbidity was persistently higher among people with low income than among those with middle/high income, and in 2018 the difference between Black and White people was not significant when stratified by income (Table 2 and S4 Fig).

**Table 2.** Change in the Adjusted Multimorbidity Prevalence from 1999 to 2018, by Race and Ethnicity.

|  | **Asian**<br>Percentage points<br>(95% CI), p value | **Black**<br>Percentage points<br>(95% CI), p value | **Latino/Hispanic**<br>Percentage points<br>(95% CI), p value | **White**<br>Percentage points<br>(95% CI), p value |
| --- | --- | --- | --- | --- |
| **Annualized rate of change in prevalence** |  |  |  |  |
| Overall | +0.23 (+0.14, +0.31),<br><0.001 | +0.31 (+0.20, +0.43),<br><0.001 | +0.31 (+0.23, +0.40),<br><0.001 | +0.28 (+0.21, +0.34),<br><0.001 |
| Low income | +0.31 (+0.15, +0.46),<br>0.001 | +0.38 (+0.23, +0.52),<br><0.001 | +0.36 (+0.27, +0.45),<br><0.001 | +0.45 (+0.34, +0.56),<br><0.001 |
| Middle/high income | +0.18 (+0.10, +0.27),<br>0.001 | +0.24 (+0.12, +0.36),<br>0.001 | +0.27 (+0.16, +0.38),<br><0.001 | +0.22 (+0.16, +0.27),<br><0.001 |
| **Absolute change in prevalence from 1999 to 2018** |  |  |  |  |
| Overall | +6.19 (+3.39, +9.00),<br><0.001 | +4.09 (+1.62, +6.55),<br>0.001 | +6.19 (+4.08, +8.30),<br><0.001 | +5.50 (+4.64, +6.36),<br><0.001 |
| Low income | +8.17 (+1.59, +14.8),<br>0.02 | +5.65 (+1.66, +9.64),<br>0.005 | +7.99 (+4.76, +11.22),<br><0.001 | +8.57 (+6.45, +10.70),<br><0.001 |
| Middle/high income | +5.68 (+2.46, +8.90),<br><0.001 | +3.33 (+0.25, +6.40),<br>0.03 | +4.88 (+1.99, +7.77),<br><0.001 | +4.70 (+3.80, +5.60),<br><0.001 |
| **Difference with White in 1999** |  |  |  |  |
| Overall | -7.32 (-9.27, -5.38),<br><0.001 | +3.95 (+2.34, +5.56),<br><0.001 | -2.76 (-4.03, -1.49),<br><0.001 | — |
| Low income | -11.06 (-15.60, -6.52), | +2.94 (+0.20, +5.68), | -6.67 (-8.76, -4.58), | — |
|  | <0.001 | 0.04 | <0.001 |  |
| Middle/high<br>income | -6.34 (-8.70, -3.97),<br><0.001 | +1.89 (-0.29, +4.06),<br>0.09 | -2.64 (-4.39, -0.89),<br>0.003 | — |
| **Difference with White**<br>**in 2018** |  |  |  |  |
| Overall | -6.63 (-8.83, -4.43),<br><0.001 | +2.54 (+0.48, +4.59),<br>0.02 | -2.07 (-3.96, -0.18),<br>0.03 | — |
| Low income | -11.46 (-16.68, -6.23),<br><0.001 | +0.02 (-3.58, +3.62),<br>0.99 | -7.25 (-10.50, -4.00),<br><0.001 | — |
| Middle/high<br>income | -5.35 (-7.72, -2.98),<br><0.001 | +0.51 (-1.85, +2.87),<br>0.67 | -2.46 (-4.92, +0.01),<br>0.05 | — |
| **Absolute change in**<br>**difference with White**<br>**from 1999 to 2018** |  |  |  |  |
| Overall | +0.69 (-2.24, +3.63),<br>0.65 | -1.41 (-4.03, +1.20),<br>0.29 | +0.69 (-1.59, +2.97),<br>0.55 | — |
| Low income | -0.40 (-7.32, +6.52),<br>0.91 | -2.92 (-7.44, +1.60),<br>0.21 | -0.58 (-4.44, +3.29),<br>0.77 | — |
| Middle/high<br>income | +0.98 (-2.36, +4.33),<br>0.57 | -1.37 (-4.58, +1.83),<br>0.40 | +0.18 (-2.84, +3.21),<br>0.91 | — |

Data source is the National Health Interview Survey from 1999 to 2018. For change in multimorbidity prevalence and change in difference: a positive sign (+) means the prevalence of multimorbidity, or its difference with White people, increased; a negative sign (-) means it decreased. The estimated multimorbidity prevalence was adjusted by age, sex, and region.
Abbreviations: CI, confidence interval

**Fig 1.**
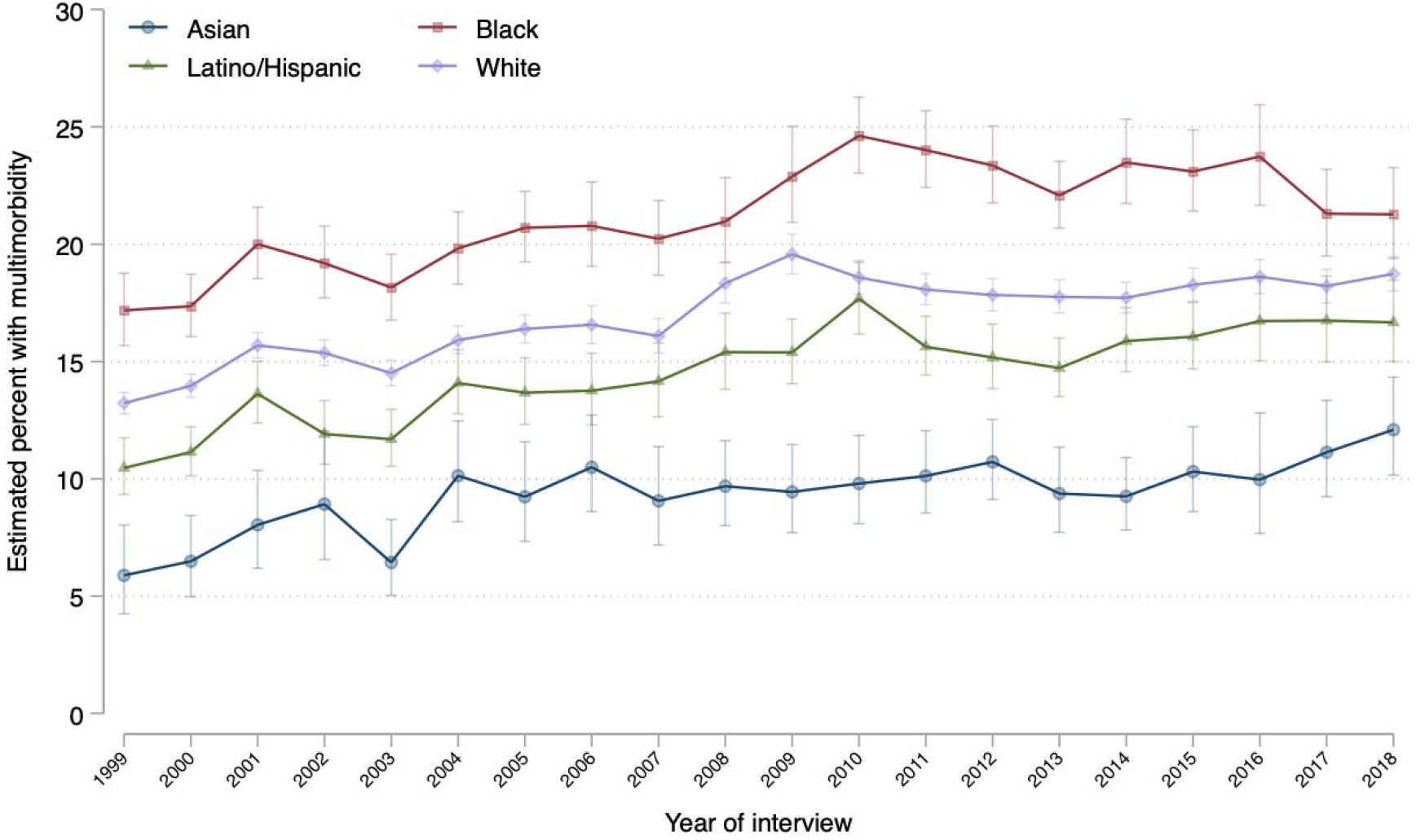
Trends in Adjusted Multimorbidity Prevalence by Race and Ethnicity, 1999–2018. Data source is the National Health Interview Survey from 1999 to 2018. Multimorbidity was defined as the presence of 2 or more of the following conditions: asthma, cancer, chronic obstructive pulmonary disease, diabetes, heart disease, hypertension, stroke, liver disease, and “weak or failing kidneys.” Annual prevalence for each race and ethnicity was obtained using logistic regression models adjusted by age, sex, and US region. The 95% confidence intervals are represented in brackets.

The estimated mean number of chronic conditions were the highest among Black individuals throughout the study period (Fig 2A). Similarly, Black individuals had the greatest prevalence of at least 1, 2, 3, or 4 conditions over the study period (Fig 2B).

**Fig 2.**
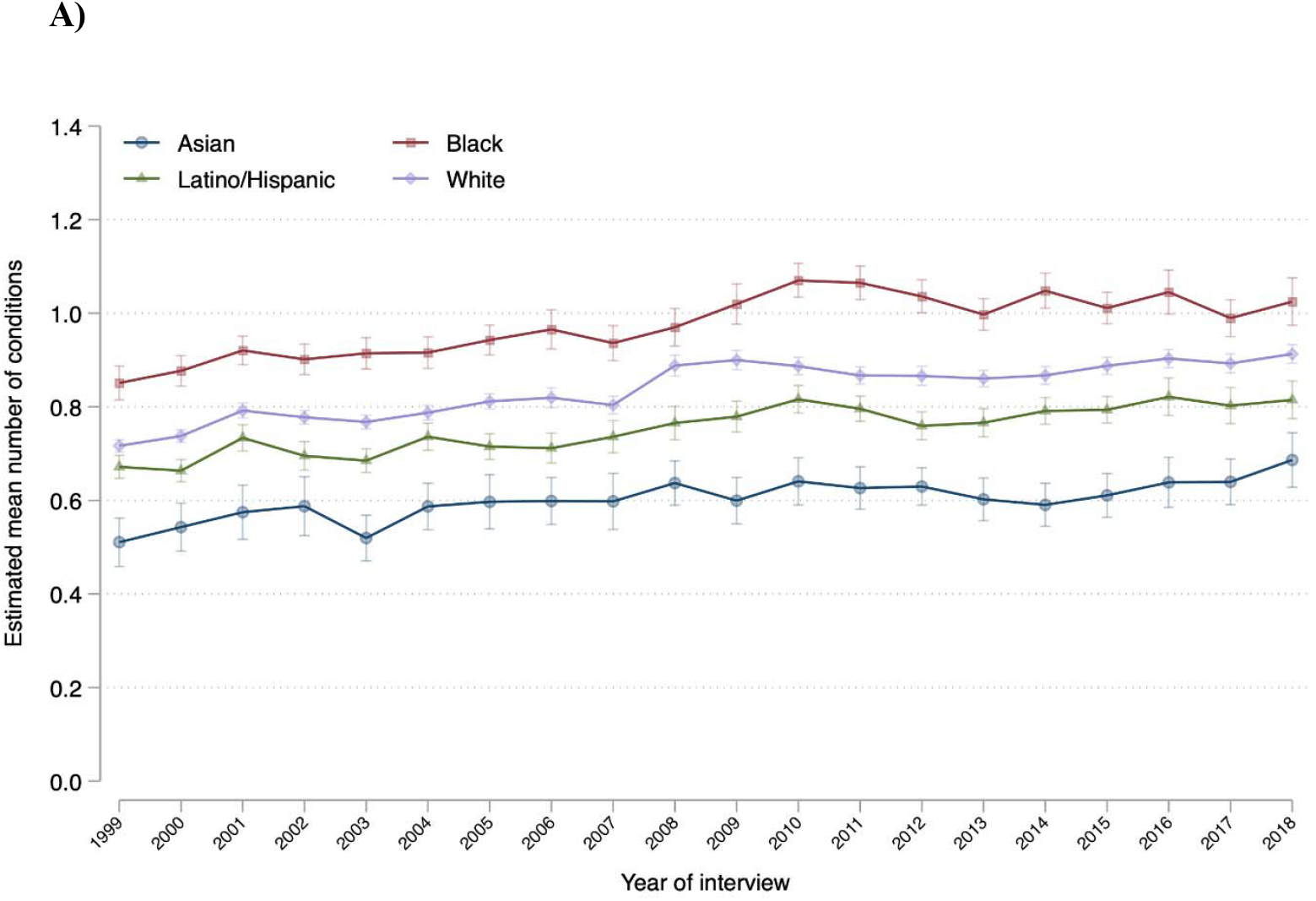
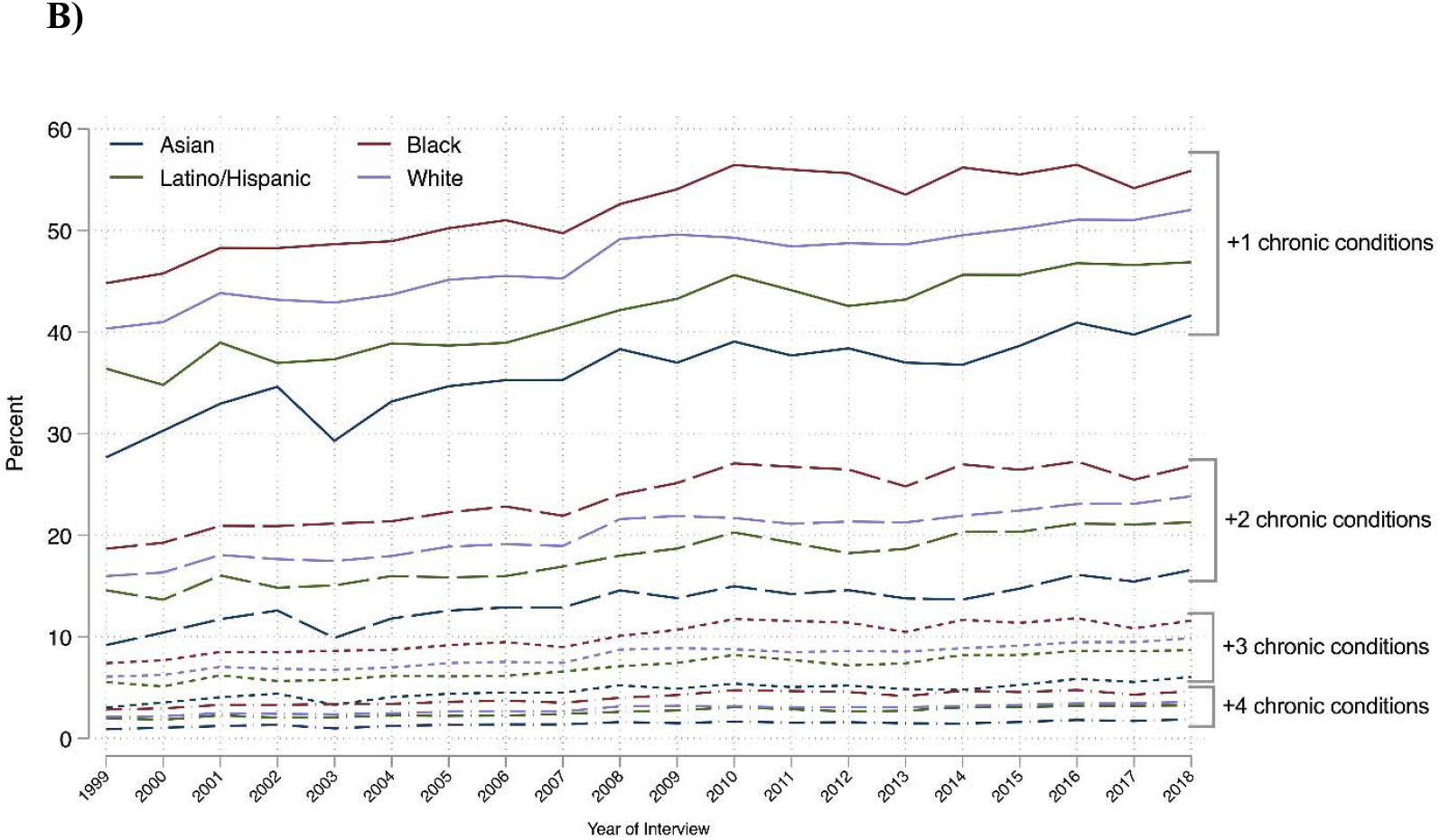
Trends in Estimated Mean Number of Chronic Conditions (A) and Ordered Number of Chronic Conditions (B) by Race and Ethnicity, 1999–2018. Data source is the National Health Interview Survey from 1999 to 2018. The mean number of chronic conditions was estimated using linear regression. The ordered number of chronic conditions was estimated using ordered logistic regression. All estimates were adjusted by age, sex, and US region. The 95% confidence intervals are represented in brackets in Panel A. For visualization purposes, no symbols or brackets are shown in panel B. Details in Methods section.

### Association Between Multimorbidity and Age

The estimated prevalence of reporting at least 1, 2, 3, or 4 conditions all increased with age across all groups, with Black people having the greatest prevalence in each age category (S5 Fig). Among those aged ≥30 years, Black people had rates of multimorbidity equivalent to those of Latino/Hispanic and White people aged 5 years older, and to those of Asian people aged 10 years older (Fig 3A). Notably, the adjusted prevalence of multimorbidity (≥2 concurrent conditions) diverged between Black people and White people among those aged 30–34 years, reached its maximum difference of 10.1 percentage points (95% CI: 8.2, 12.1) among those aged 60–64, and converged to reach similar rates among those aged ≥80 (Fig 3B).

**Fig 3.**
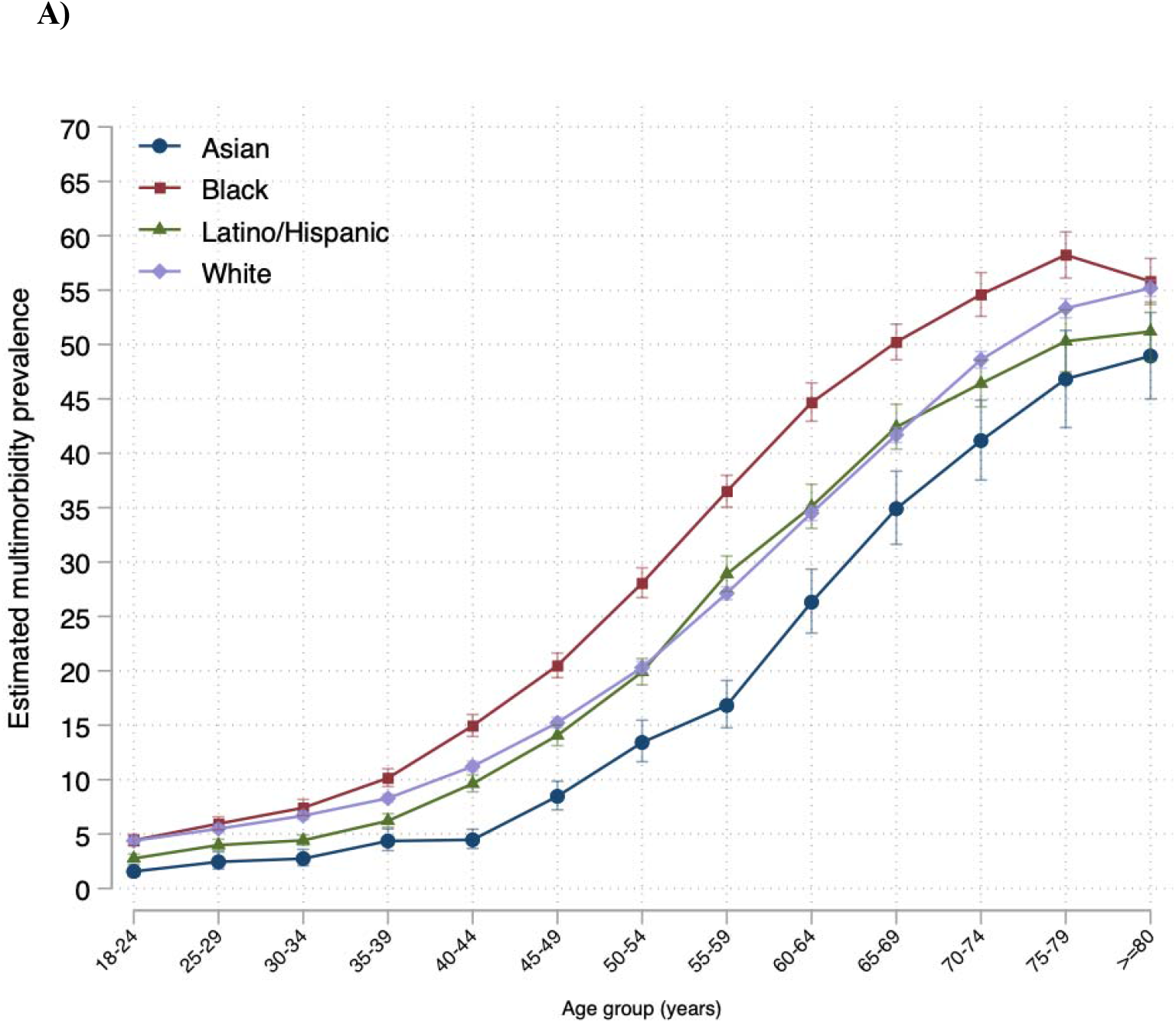
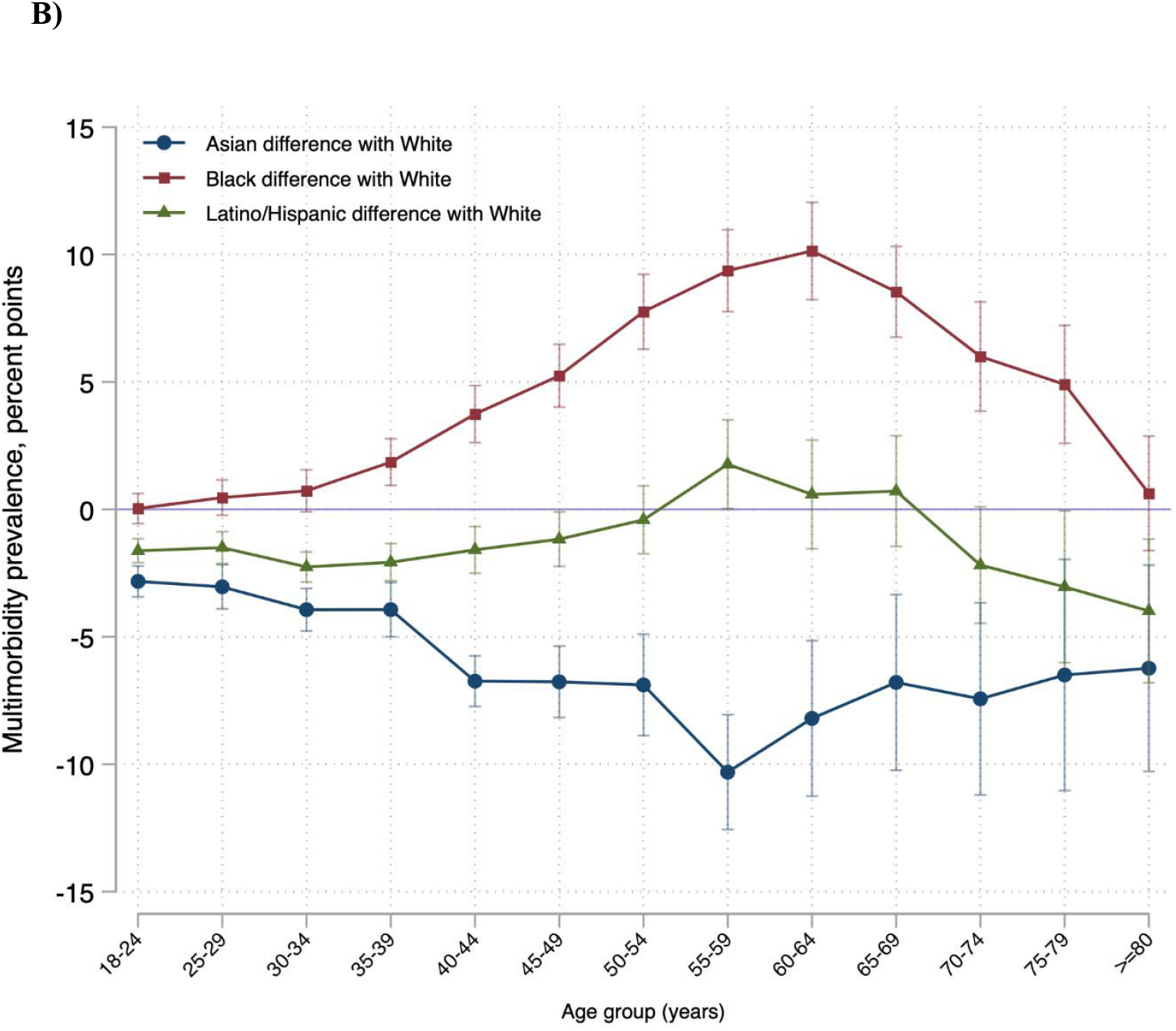
Adjusted Multimorbidity Prevalence by Age and Race and Ethnicity (A), and Prevalence Differences by Age and Race and Ethnicity (B). Data source is the National Health Interview Survey from 1999 to 2018. Panel A shows the sex- and region-adjusted prevalence of multimorbidity by age for each race and ethnicity group. Panel B shows the difference in such a prevalence between Asian, Black, and Latino/Hispanic people with White people of the same age group (e.g., prevalence of multimorbidity among Black people aged 50–54 years - prevalence of multimorbidity among White people aged 50–54 years). The 95% confidence intervals are represented in brackets.

There were significant racial and ethnic differences in the association between age and prevalence of multimorbidity. Asian, Black, and Latino/Hispanic people had a greater increase in the prevalence of multimorbidity with age compared with White people (multimorbidity*decade of age interaction term OR 1.17 [95% CI: 1.13, 1.20] for Asian, OR 1.04 [95% CI: 1.02, 1.06] for Black, and OR 1.10 [95% CI: 1.08, 1.12] for Latino/Hispanic; P<0.001 for each). Within each racial and ethnic group, people with low income were associated with a higher prevalence of multimorbidity compared with people in the same age group who had middle/high income (S6 Fig).

### Sensitivity Analysis

Results from our sensitivity analysis, including arthritis in the chronic conditions count (in the years it was available), were mostly consistent with our main results, with some notable differences (S8-11 Figs). Prevalence of multimorbidity, as expected, was higher. From 2002 to 2018, the estimated prevalence of multimorbidity was 27.2% (95% CI: 27.0%, 27.5%). In this sensitivity analysis, the difference in the prevalence of multimorbidity between Black and White people decreased significantly from 2002 to 2018 and was not significant in 2018 (S8 Fig).

## DISCUSSION

From 1999 to 2018, there were increasing rates of multimorbidity and a static racial gap in which Black people persistently had the highest prevalence and Asian people had the lowest. Moreover, multimorbidity prevalence was higher among Black people compared with Asian, Latino/Hispanic, and White people of the same age range, with differences emerging among those 30–34 years old and peaking among those 60–64 years old. Overall, and within people of the same racial and ethnic group, those with low income experienced a higher burden of multimorbidity. This 20-year period, which corresponded with significant health care investments and dramatic increases in per capita health care costs,[21, 22] fails to demonstrate meaningful progress in reducing multimorbidity or racial and ethnic disparities in its prevalence.

Studies have reported racial/ethnic disparities but, to the best of our knowledge, this is the first to analyze whether there was improvement during a recent period notable for increased national attention on reducing health disparities. Our findings expand the literature in several ways. First, we found no improvement in the race/ethnicity gap in multimorbidity. The increasing prevalence of multimorbidity occurred almost in parallel across each of the racial and ethnic groups. These findings reveal a lack of progress in narrowing racial/ethnic disparities in health over a 20-year period, in which Black individuals persistently had the highest multimorbidity prevalence despite major nationwide policies aimed to address health disparities.[23-26] Notably, multimorbidity was consistently less prevalent among Asian individuals, even when stratified by household income level. Although unclear, such a lower chronic conditions burden may be due to generally more favorable socioeconomic conditions among Asian individuals, including higher educational level, that may facilitate resources to maintain healthy habits and a healthy life.[27] Nonetheless, this finding may also represent an underestimation of the true multimorbidity prevalence among Asian people due to barriers to health care access and utilization that may prevent the detection of chronic conditions.[28] Studies designed to better understand this pattern are needed.

Second, we found that the racial and ethnic gap was concentrated among non-elderly adults (<65 years old), emerging among those aged 30–34 years, and peaking among those 60– 64 years old. It is unclear if such age-related divergence in multimorbidity prevalence may be due to a cumulative effect of an earlier life concentration of risk factors that Black people are more likely to be exposed to, such as environmental stressors, smoking, and barriers to health care.[11, 29, 30] The convergence among the elderly, on the other hand, may be explained by a combination of a ceiling effect in chronic conditions and differences in risk of death associated with earlier multimorbidity among Black people.[31]

Third, we found that stratification by income level attenuated the disparities. This observation underscores the role of income inequality in the perpetuation of racial and ethnic disparities in health.[32] This finding is especially important given the disproportionately large percentage of Black people in the low-income stratum[33] group that during the study period also reported the highest prevalence of poor or fair health.[11] Further studies are required to better understand the association between income (or other sociodemographic characteristics such as education and occupation) and multimorbidity, and how such associations may have changed over the years.

There are some differences between our findings and the results from other studies that have measured multimorbidity using data from the NHIS. First, the overall unadjusted weighted prevalence of multimorbidity in our study (19.9%) was lower than the estimates from others.[17-19] This discrepancy is likely due to the inclusion of arthritis in their calculation of multimorbidity, which was inconsistently available across the 20 years of this study. In an exploratory estimation of multimorbidity using data from 2002 to 2018 including arthritis, the overall estimates were consistent with those from previous reports and did not affect the overall findings. Second, our annual estimates of multimorbidity were adjusted for age, sex, and region, which can provide greater insight into disparities by race and ethnicity. Of particular importance is the higher mean age among White individuals when compared with Black individuals which, considering the strong association between age and multimorbidity, if left unaccounted for would underestimate the gap between the two groups.

This study has important implications. First, it shows that the public health challenge of multimorbidity is increasing among the 4 largest race and ethnicity groups in the US, underscoring the growing need for preventive measures nationwide. People with multimorbidity experience challenges not only related to their individual conditions but also related to the complexity and costs of managing multiple conditions simultaneously. Multimorbidity is associated with worse health status,[34] faster functional decline,[35] lower quality of life,[34, 36] higher mortality,[37] higher rates of unmet medical needs,[38] and higher health care costs,[39, 40] and this study shows that the burden has been persistently higher among Black people.

Although preventing the development of multimorbidity should be a public health priority, these findings suggest that complementary and targeted strategies are needed for Black people. The observed persistent gap in multimorbidity over the study period may be associated with the persistent disparity in access to preventive health care services that Black people experience. In 2018, and despite modest improvements since 1999, Black people were more likely to lack health insurance coverage, to lack a usual source of health care, and to report unmet medical needs due to cost compared with White people.[11] Also, stressors derived from race-based discrimination are associated with higher burden of chronic diseases and multimorbidity.[41, 42] Thus, our findings should be interpreted in the context of vast literature that identifies systemic racism as a fundamental determinant of health inequalities through the reinforcement of an artificial racial and ethnic hierarchy that puts Black people at a disadvantage in obtaining access to goods and opportunities necessary for a healthy life. Public health strategies should account for this in the distribution of resources and efforts to prevent the development of chronic conditions over a lifetime. Particularly, the emergence of the disparities among young adults suggests that such strategies must aim for the prevention of the onset of chronic conditions early in life.

Our study has several limitations. First, the cross-sectional design prevents us from measuring and comparing the incidence of multimorbidity across the lifetime of an individual by racial and ethnic groups. Furthermore, when analyzing the differences with age, the study design may be introducing survivor bias as age group increases, which may underestimate the overall impact of multimorbidity on health disparities. Second, based on the US Department of Health and Human Services conceptual framework for studying multiple chronic conditions, we used a limited number of chronic conditions based on their availability in the NHIS dataset, but lacked information on the severity, complexity, duration, and status of such conditions. Also importantly, none of these were mental health conditions. Third, our multimorbidity measure may be an underestimation because the study relied on self-reported conditions that may go underreported due to barriers to access to care—for its diagnosis—and low health literacy. If any, such underreporting more likely affects Black and Latino/Hispanic people because of their higher barriers to health care. Finally, we focused on large racial and ethnic group categories and were underpowered to examine a wider set of racial and ethnic groups.

### Conclusions

From 1999 to 2018, multimorbidity in the US increased and Black people persistently had the highest prevalence of multimorbidity, with racial gaps that did not change. When analyzed by age, this gap emerged among young adults and peaked among those aged 60–64 years. Public health interventions that prevent the onset of chronic conditions in early life may be needed to eliminate these disparities.

## Data Availability

Data from the National Health Interview Survey is publicly available. We obtained the data from the Integrated Public Use Microdata Series Health Surveys (https://nhis.ipums.org). The code used to in the analysis is publicly available at http://doi.org/10.3886/E151983V1.

## ACKNOWLEDGMENTS

N/A

## SOURCE OF FUNDING

The study was self-funded.

## COMPETING INTERESTS DISCLOSURE

All authors have completed the ICMJE uniform disclosure form at www.icmje.org/coi_disclosure.pdf and declare: In the past three years, **Harlan Krumholz** received expenses and/or personal fees from UnitedHealth, IBM Watson Health, Element Science, Aetna, Facebook, the Siegfried and Jensen Law Firm, Arnold and Porter Law Firm, Martin/Baughman Law Firm, F-Prime, and the National Center for Cardiovascular Diseases in Beijing. He is a co-founder of Refactor Health and HugoHealth, and had grants and/or contracts from the Centers for Medicare & Medicaid Services, Johnson & Johnson, Foundation for a Smoke-Free World, State of Connecticut Department of Public Health, Agency for Healthcare Research and Quality, American Heart Association, and the National Institutes of Health. **Yuan Lu** is supported by the National Heart, Lung, and Blood Institute (K12HL138037) and the Yale Center for Implementation Science. **Jeph Herrin** has had grants and/or contracts from the Centers for Medicare & Medicaid Services, the National Institutes of Health, Mayo Clinic, Yale University, and Mercy Health. **Chima Ndumele** has had grants from the National Institutes of Health, Robert Wood Johnson Foundation, and the Commonwealth Fund. Other authors declare no competing interests.

## AUTHORS CONTRIBUTIONS

HK conceived the study and supervised the manuscript preparation and data interpretation. CC and SM obtained the data. CC performed the literature review and analysis, and wrote the first draft of the manuscript. JH supervised and verified the statistical methods. CC, DM, SM, YL, NC, ED, JH, and HK interpreted the results. SM, DM, YL, NC, ED, JH, and HK critically reviewed the manuscript, and made significant intellectual contributions to the study. The corresponding author attests that all listed authors meet authorship criteria and that no others meeting the criteria have been omitted.

## SUPPORTING INFORMATION

**Supporting Information Methods: Statistical Analysis**.

**S1 Table. Study Population Characteristics**.

**S1 Fig. Study Population Flowchart**.

**S2 Fig. Unadjusted Age Distribution by Race and Ethnicity**.

**S3 Fig. Unadjusted Distribution of Number of Chronic Conditions by Race and Ethnicity**

**S4 Fig. Trends in Adjusted Prevalence of Individual Conditions by Race and Ethnicity, 1999-2018**.

**S5 Fig. Trends in Adjusted Multimorbidity Prevalence by Race, Ethnicity, and Income (A) and by Income Alone (B), 1999-2018**.

**S6 Fig. Number of Chronic Conditions by Age Among Asian, Black, Latino/Hispanic, and White People**.

**S7 Fig. Adjusted Multimorbidity Prevalence by Age, Race, Ethnicity, and Income (A), and by Age and Income (B)**.

**S8 Fig. Sensitivity Analysis Including Arthritis: Trends in Adjusted Multimorbidity Prevalence by Race and Ethnicity, 2002-2018**.

**S9 Fig. Sensitivity Analysis Including Arthritis: Number of Chronic Conditions by Age Among Asian, Black, Latino/Hispanic, and White People**.

**S10 Fig. Sensitivity Analysis Including Arthritis: Adjusted Multimorbidity Prevalence by Age, Race, and Ethnicity (A), and Prevalence Differences by Age, Race, and Ethnicity (B)**.

**S11 Fig. Sensitivity Analysis: Trends in Adjusted Prevalence of Arthritis by Race and Ethnicity, 2002-2018**.

## Notes

### Funding Statement

This study was self-funded

### Author Declarations

The Institutional Review Board at Yale University exempted the study from review.

## REFERENCES

1. US Department of Health Human Services. Multiple chronic conditions—a strategic framework: optimum health and quality of life for individuals with multiple chronic conditions. Washington, DC: US Department of Health and Human Services. 2010;2.

2. Goodman RA, Posner SF, Huang ES, Parekh AK, Koh HK. Defining and measuring chronic conditions: imperatives for research, policy, program, and practice. Prev Chronic Dis. 2013;10:E66. Epub 2013/04/27. doi: 10.5888/pcd10.120239. PubMed PMID: 23618546; PubMed Central PMCID: PMCPMC3652713.

3. King DE, Xiang J, Pilkerton CS. Multimorbidity Trends in United States Adults, 1988-2014. J Am Board Fam Med. 2018;31(4):503–13. Epub 2018/07/11. doi: 10.3122/jabfm.2018.04.180008. PubMed PMID: 29986975; PubMed Central PMCID: PMCPMC6368177.

4. Johnson-Lawrence V, Zajacova A, Sneed R. Education, race/ethnicity, and multimorbidity among adults aged 30-64 in the National Health Interview Survey. SSM Popul Health. 2017;3:366–72. Epub 2018/01/20. doi: 10.1016/j.ssmph.2017.03.007. PubMed PMID: 29349230; PubMed Central PMCID: PMCPMC5769041.

5. Rocca WA, Boyd CM, Grossardt BR, Bobo WV, Finney Rutten LJ, Roger VL, et al. Prevalence of multimorbidity in a geographically defined American population: patterns by age, sex, and race/ethnicity. Mayo Clin Proc. 2014;89(10):1336–49. Epub 2014/09/16. doi: 10.1016/j.mayocp.2014.07.010. PubMed PMID: 25220409; PubMed Central PMCID: PMCPMC4186914.

6. Centers for Disease Control & Prevention (CDC). About the National Health Interview Survey: National Center for Health Statistics; [cited 2021 November 26]. Available from: https://www.cdc.gov/nchs/nhis/about_nhis.htm.

7. Blewett LA, Rivera Drew JA, King ML, and Williams KCW. IPUMS health surveys: National Health Interview Survey, version 6.4 [dataset]. Minneapolis, MN: IPUMS, 2019. [cited 2021 November 26]. Available from: https://doi.org/10.18128/D070.V6.4.

8. United States Census Bureau. People in families by family structure, age, and sex, iterated by income-to-poverty ratio and race [cited 2021 November 26]. Available from: https://www.census.gov/data/tables/time-series/demo/income-poverty/cps-pov/pov-02.html#par_textimage_30.

9. Caraballo C, Valero-Elizondo J, Khera R, Mahajan S, Grandhi GR, Virani SS, et al. Burden and consequences of financial hardship from medical bills among nonelderly adults with diabetes mellitus in the United States. Circ Cardiovasc Qual Outcomes. 2020;13(2):e006139. Epub 2020/02/19. doi: 10.1161/circoutcomes.119.006139. PubMed PMID: 32069093.

10. Dubay LC, Lebrun LA. Health, behavior, and health care disparities: disentangling the effects of income and race in the United States. Int J Health Serv. 2012;42(4):607–25. Epub 2013/02/02. doi: 10.2190/HS.42.4.c. PubMed PMID: 23367796.

11. Mahajan S, Caraballo C, Lu Y, Valero-Elizondo J, Massey D, Annapureddy AR, et al. Trends in differences in health status and health care access and affordability by race and ethnicity in the United States, 1999-2018. JAMA. 2021;326(7):637–48. Epub 2021/08/18. doi: 10.1001/jama.2021.9907. PubMed PMID: 34402830.

12. Drye EE, Altaf FK, Lipska KJ, Spatz ES, Montague JA, Bao H, et al. Defining multiple chronic conditions for quality measurement. Med Care. 2018;56(2):193–201.

13. National Quality Forum. Multiple chronic conditions measurement framework. 2012 [cited 2021 November 20]. Available from: https://www.qualityforum.org/Publications/2012/05/MCC_Measurement_Framework_Final_Report.aspx.

14. Boyd CM, Fortin M. Future of multimorbidity research: how should understanding of multimorbidity inform health system design? Public Health Reviews. 2010;32(2):451–74.

15. Division of Health Interview Statistics National Center for Health Statistics. Multiple imputation of family income and personal earnings in the National Health Interview Survey: methods and examples. August 2019 https://nhis.ipums.org/nhis/resources/tecdoc18.pdf [cited 2021 November 16]. Available from: https://nhis.ipums.org/nhis/resources/tecdoc18.pdf.

16. Barnett K, Mercer SW, Norbury M, Watt G, Wyke S, Guthrie B. Epidemiology of multimorbidity and implications for health care, research, and medical education: a cross-sectional study. The Lancet. 2012;380(9836):37–43.

17. Boersma P, Black LI, Ward BW. Prevalence of multiple chronic conditions among US adults, 2018. Prev Chronic Dis. 2020;17:E106. Epub 2020/09/19. doi: 10.5888/pcd17.200130. PubMed PMID: 32945769; PubMed Central PMCID: PMCPMC7553211.

18. Ward BW, Schiller JS, Goodman RA. Peer reviewed: multiple chronic conditions among US adults: a 2012 update. Preventing Chronic Disease. 2014;11.

19. Ward BW, Schiller JS. Prevalence of multiple chronic conditions among US adults: estimates from the National Health Interview Survey, 2010. Prev Chronic Dis. 2013;10:E65. Epub 2013/04/27. doi: 10.5888/pcd10.120203. PubMed PMID: 23618545; PubMed Central PMCID: PMCPMC3652717.

20. Centers for Disease Control and Prevention. National Health Interview Survey, 1997-2018. Survey description document. [cited 2021 November 30]. Available from: https://www.cdc.gov/nchs/nhis/1997-2018.htm.

21. Centers for Medicare & Medicaid Services. National Health Expenditure Data, Historical. December 2019. [cited 2021 November 17] Available from: https://www.cms.gov/Research-Statistics-Data-and-Systems/Statistics-Trends-and-Reports/NationalHealthExpendData/NationalHealthAccountsHistorical.

22. Kamal R, McDermott D, Cox C. Peterson Center on Healthcare-Kaiser Family Foundation Health System Tracker. How has U.S. spending on healthcare changed over time? December 2019 [cited 2021 November 26]. Available from: https://www.healthsystemtracker.org/chart-collection/u-s-spending-healthcare-changed-time/.

23. United States Department of Health and Human Services. Task Force on Black and Minority Health. Report of the Secretary’s Task Force on Black and Minority Health. 1985. [cited 2021 November 26]. Available from: http://resource.nlm.nih.gov/8602912.

24. U.S. Department of Health and Human Services. About the Office of Minority Health https://www.minorityhealth.hhs.gov/omh/browse.aspx?lvl=1&lvlid=1 x[cited 2021 November 16]. Available from https://www.minorityhealth.hhs.gov/omh/browse.aspx?lvl=1&lvlid=1.

25. Koh HK, Graham G, Glied SA. Reducing racial and ethnic disparities: the action plan from the Department of Health and Human Services. Health Aff (Millwood). 2011;30(10):1822–9. Epub 2011/10/07. doi: 10.1377/hlthaff.2011.0673. PubMed PMID: 21976322.

26. U.S. Department of Health and Human Services. Healthy People. Foundation health measures archive: disparities [cited 2021 November 16]. Available from: https://www.healthypeople.gov/2020/about/foundation-health-measures/Disparities.

27. Zhang Y, Misra R, Sambamoorthi U. Prevalence of multimorbidity among Asian Indian, Chinese, and Non-Hispanic White adults in the United States. International Journal of Environmental Research and Public Health. 2020;17(9):3336. PubMed PMID: doi:10.3390/ijerph17093336.

28. Lee S, Martinez G, Ma GX, Hsu CE, Robinson ES, Bawa J, et al. Barriers to health care access in 13 Asian American communities. Am J Health Behavior. 2010;34(1):21–30.

29. Bailey ZD, Krieger N, Agénor M, Graves J, Linos N, Bassett MT. Structural racism and health inequities in the USA: evidence and interventions. The Lancet. 2017;389(10077):1453–63.

30. Bailey ZD, Feldman JM, Bassett MT. How structural racism works — racist policies as a root cause of U.S. racial health inequities. N Engl J Med. 2020;384(8):768–73. doi: 10.1056/NEJMms2025396.

31. Quiñones AR, Newsom JT, Elman MR, Markwardt S, Nagel CL, Dorr DA, et al. Racial and ethnic differences in multimorbidity changes over time. Med Care. 2021;59(5):402–9. Epub 2021/04/07. doi: 10.1097/mlr.0000000000001527. PubMed PMID: 33821829; PubMed Central PMCID: PMCPMC8024615.

32. Bor J, Cohen GH, Galea S. Population health in an era of rising income inequality: USA, 1980-2015. The Lancet. 2017;389(10077):1475–90. doi: 10.1016/S0140-6736(17)30571-8.

33. Bhutta N, Chang AC, Dettling LJ, Hsu JW. Disparities in wealth by race and ethnicity in the 2019 Survey of Consumer Finances. FEDS Notes. Washington: Board of Governors of the Federal Reserve System, September 28, 2020. [cited 2021 November 16]. Available from: https://www.federalreserve.gov/econres/notes/feds-notes/disparities-in-wealth-by-race-and-ethnicity-in-the-2019-survey-of-consumer-finances-20200928.htm.

34. Arokiasamy P, Uttamacharya U, Jain K, Biritwum RB, Yawson AE, Wu F, et al. The impact of multimorbidity on adult physical and mental health in low-and middle-income countries: what does the Study on Global Ageing and Adult Health (SAGE) reveal? BMC Med. 2015;13:178. Epub 2015/08/05. doi: 10.1186/s12916-015-0402-8. PubMed PMID: 26239481; PubMed Central PMCID: PMCPMC4524360.

35. Ryan A, Wallace E, O’Hara P, Smith SM. Multimorbidity and functional decline in community-dwelling adults: a systematic review. Health Qual Life Outcomes. 2015;13:168. Epub 2015/10/16. doi: 10.1186/s12955-015-0355-9. PubMed PMID: 26467295; PubMed Central PMCID: PMCPMC4606907.

36. Kanesarajah J, Waller M, Whitty JA, Mishra GD. Multimorbidity and quality of life at mid-life: A systematic review of general population studies. Maturitas. 2018;109:53–62. Epub 2018/02/18. doi: 10.1016/j.maturitas.2017.12.004. PubMed PMID: 29452782.

37. Nunes BP, Flores TR, Mielke GI, Thumé E, Facchini LA. Multimorbidity and mortality in older adults: A systematic review and meta-analysis. Arch Gerontol Geriatr. 2016;67:130–8. Epub 2016/08/09. doi: 10.1016/j.archger.2016.07.008. PubMed PMID: 27500661.

38. Johnson PJ, Mentzer KM, Jou J, Upchurch DM. Unmet healthcare needs among midlife adults with mental distress and multiple chronic conditions. Aging Ment Health. 2021:1–9. Epub 2021/04/02. doi: 10.1080/13607863.2021.1904830. PubMed PMID: 33792432.

39. Lee JT, Hamid F, Pati S, Atun R, Millett C. Impact of noncommunicable disease multimorbidity on healthcare utilisation and out-of-pocket expenditures in middle-income countries: cross sectional analysis. PLoS One. 2015;10(7):e0127199. Epub 2015/07/15. doi: 10.1371/journal.pone.0127199. PubMed PMID: 26154083; PubMed Central PMCID: PMCPMC4496037.

40. Wang L, Si L, Cocker F, Palmer AJ, Sanderson K. A systematic review of cost-of-illness studies of multimorbidity. Appl Health Econ Health Policy. 2018;16(1):15–29. Epub 2017/09/01. doi: 10.1007/s40258-017-0346-6. PubMed PMID: 28856585.

41. Mays VM, Cochran SD, Barnes NW. Race, Race-based discrimination, and health outcomes among African Americans. Ann Rev Psychol. 2007;58(1):201–25. doi: 10.1146/annurev.psych.57.102904.190212. PubMed PMID: 16953796.

42. Oh H, Glass J, Narita Z, Koyanagi A, Sinha S, Jacob L. Discrimination and multimorbidity among black Americans: Findings from the National Survey of American Life. J Racial Ethnic Health Disparities. 2021;8(1):210–9. doi: 10.1007/s40615-020-00773-z.

