## Supporting Information for "Temporal Trends in Racial and Ethnic Disparities in Multimorbidity Prevalence in the United States, 1999–2018"

### TABLE OF CONTENTS


**Supporting Information Methods: Statistical Analysis.**

**S1 Table. Study Population Characteristics.**

**S1 Figure. Study Population Flowchart.**

**S2 Figure. Unadjusted Age Distribution by Race and Ethnicity.**

**S3 Figure. Unadjusted Distribution of Number of Chronic Conditions by Race and Ethnicity**

**S4 Figure. Trends in Adjusted Prevalence of Individual Conditions by Race and Ethnicity, 1999-2018.**

**S5 Figure. Trends in Adjusted Multimorbidity Prevalence by Race, Ethnicity, and Income (A) and by Income Alone (B), 1999-2018.**

**S6 Figure. Number of Chronic Conditions by Age Among Asian, Black, Latino/Hispanic, and White People.**

**S7 Figure. Adjusted Multimorbidity Prevalence by Age, Race, Ethnicity, and Income (A), and by Age and Income (B).**

**S8 Figure. Sensitivity Analysis Including Arthritis: Trends in Adjusted Multimorbidity Prevalence by Race and Ethnicity, 2002-2018.**

**S9 Figure. Sensitivity Analysis Including Arthritis: Number of Chronic Conditions by Age Among Asian, Black, Latino/Hispanic, and White People.**

**S10 Figure. Sensitivity Analysis Including Arthritis: Adjusted Multimorbidity Prevalence by Age, Race, and Ethnicity (A), and Prevalence Differences by Age, Race, and Ethnicity (B).**

**S11 Figure. Sensitivity Analysis: Trends in Adjusted Prevalence of Arthritis by Race and Ethnicity, 2002-2018.**

**Supporting Information Methods: Statistical Analysis.**

The annual multimorbidity prevalence for each group was estimated using multivariable logistic regression models. In these models, multimorbidity was the dependent variable, and age, sex, a dummy variable for each region, and an indicator for each year of interview were the independent variables. Age, sex, and region were centered on their overall mean for the study sample; the coefficients for each year, when combined with the intercept, then represented the logit of the annual multimorbidity rates adjusted for age, sex, and region. A separate model was estimated for each racial and ethnic subgroup and the results were used to generate estimated rates for each year, using the inverse logit of each year effect as the annual rate and applying the method of parametric bootstrapping to calculate the standard error (SE) and the confidence interval (CI) for the transformed coefficients.[1]

The multimorbidity prevalence and racial/ethnic differences trends over the study period were estimated by fitting weighted linear regression models where the dependent variable was the adjusted annual multimorbidity prevalence or difference, and the independent variable was time in years. Each observation was weighted by the inverse square of the SE to account for varying precision of each estimated rate or the difference over time.

Following recommendations from the National Center for Health Statistics for multiply imputed data analysis,[2] to estimate the annual low-income prevalence by race and ethnicity we used the mean annual estimate obtained by separate logistic regressions using a similar approach as above, but with each of the multiply imputed low-income variables as the dependent variable and an indicator for each year as the independent variables. Similarly, we used the mean prevalence estimate of multimorbidity obtained from separate regressions for each of the income groups.

---

**References**

1. King G, Tomz M, Wittenberg J. Making the most of statistical analyses: Improving interpretation and presentation. Am J Pol Sci 2000:347-61.
2. National Center for Health Statistics. Multiple imputation of family income and personal earnings in the National Health Interview Survey: methods and examples. August 2019. (Accessed October 15, 2021, at https://nhis.ipums.org/nhis/resources/tecdoc18.pdf.)

### S1 Table. Study Population Characteristics.

| | Asian | Black | Latino/Hispanic | White |
|---|---|---|---|---|
| Sample size [N=596,236] | 27,755 | 84,942 | 99,650 | 383,889 |
| **Age in years** | 41 (30–55) | 42 (29–55) | 38 (28–50) | 47 (33–61) |
| **Age category** | | | | |
| 18–39 years | 45.7 (44.7 to 46.6) | 45.4 (44.8 to 46.0) | 54.2 (53.7 to 54.8) | 35.3 (35.0 to 35.7) |
| 40–64 years | 41.5 (40.7 to 42.4) | 41.8 (41.3 to 42.3) | 36.7 (36.2 to 37.1) | 44.4 (44.1 to 44.7) |
| ≥65 years | 12.8 (12.3 to 13.5) | 12.8 (12.4 to 13.2) | 9.1 (8.8 to 9.4) | 20.3 (20.0 to 20.6) |
| **Women** | 52.3 (51.6 to 53.1) | 55.2 (54.7 to 55.7) | 49.6 (49.1 to 50.0) | 51.7 (51.5 to 51.9) |
| **US Citizenship** [n=594,859] | 68.1 (67.0 to 69.1) | 95.3 (95.0 to 95.7) | 64.4 (63.6 to 65.2) | 98.4 (98.3 to 98.5) |
| **Education level** [n=591,667] | | | | |
| Less than high school | 9.9 (9.3 to 10.5) | 18.3 (17.8 to 18.9) | 36.9 (36.2 to 37.6) | 10.3 (10.1 to 10.5) |
| High school diploma /GED | 16.4 (15.7 to 17.1) | 30.6 (30.1 to 31.1) | 26.2 (25.8 to 26.7) | 27.9 (27.6 to 28.2) |
| Some college | 22.4 (21.7 to 23.2) | 32.7 (32.2 to 33.3) | 24.3 (23.8 to 24.8) | 31.0 (30.7 to 31.3) |
| ≥Bachelor's degree | 51.3 (50.1 to 52.5) | 18.4 (17.9 to 18.9) | 12.6 (12.2 to 13.0) | 30.8 (30.4 to 31.2) |
| **Income <200% Federal poverty level** [a] | 28.2 (24.9 to 31.7) | 46.1 (43.9 to 48.3) | 51.5 (50.2 to 52.7) | 23.9 (23.0 to 24.9) |
| **Uninsured at the time of interview** [n=594,011] | 12.9 (12.3 to 13.5) | 18.5 (18.1 to 18.9) | 34.1 (33.4 to 34.8) | 10.5 (10.4 to 10.7) |
| **US region of residence** [b] | | | | |

| | Asian | Black | Latino/Hispanic | White |
|---|---|---|---|---|
| Northeast | 20.1 (18.9 to 21.4) | 16.3 (15.5 to 17.0) | 14.0 (13.3 to 14.8) | 19.3 (18.8 to 19.8) |
| Midwest | 13.3 (12.3 to 14.3) | 17.8 (17.0 to 18.7) | 9.0 (8.3 to 9.8) | 28.2 (27.6 to 28.8) |
| South | 21.7 (20.4 to 23.0) | 57.8 (56.6 to 59.1) | 36.3 (35.0 to 37.6) | 33.9 (33.3 to 34.6) |
| West | 44.9 (43.3 to 46.6) | 8.1 (7.7 to 8.5) | 40.7 (39.3 to 42.1) | 18.6 (18.2 to 19.1) |
| **Married or living with partner** [n=593,807] | 64.5 (63.6 to 65.3) | 35.1 (34.6 to 35.7) | 53.9 (53.4 to 54.4) | 58.5 (58.1 to 58.9) |
| **Employment status** [n=595,478] | | | | |
| With a job/Working | 65.3 (64.5 to 66.2) | 60.5 (59.9 to 61.0) | 65.4 (64.9 to 65.9) | 62.9 (62.6 to 63.2) |
| Not in labor force | 30.8 (29.9 to 31.6) | 32.0 (31.4 to 32.5) | 29.3 (28.8 to 29.8) | 34.0 (33.7 to 34.3) |
| Unemployed | 3.9 (3.7 to 4.2) | 7.6 (7.3 to 7.9) | 5.3 (5.1 to 5.5) | 3.2 (3.1 to 3.2) |
| **Current smoker** | 10.2 (9.7 to 10.7) | 19.7 (19.3 to 20.2) | 13.4 (13.1 to 13.7) | 20.6 (20.4 to 20.9) |
| **Flu vaccine in past 12 months** | 36.8 (35.9 to 37.6) | 27.1 (26.7 to 27.6) | 24.6 (24.1 to 25.0) | 37.2 (36.9 to 37.5) |
| **Obese (BMI ≥30 kg/m$^2$)** | 9.1 (8.7 to 9.6) | 36.3 (35.8 to 36.8) | 29.6 (29.1 to 30.1) | 25.6 (25.4 to 25.9) |
| **Conditions** | | | | |
| Asthma | 8.0 (7.6 to 8.5) | 13.2 (12.9 to 13.5) | 9.5 (9.3 to 9.8) | 12.1 (12.0 to 12.2) |
| Cancer | 2.9 (2.7 to 3.2) | 4.0 (3.8 to 4.1) | 2.8 (2.7 to 2.9) | 10.0 (9.9 to 10.1) |
| COPD | 1.8 (1.6 to 2.0) | 4.7 (4.5 to 4.9) | 2.8 (2.7 to 2.9) | 5.9 (5.8 to 6.0) |
| Diabetes | 7.2 (6.8 to 7.6) | 11.0 (10.7 to 11.2) | 8.6 (8.3 to 8.9) | 7.7 (7.5 to 7.8) |
| Heart disease | 5.7 (5.3 to 6.0) | 9.6 (9.3 to 9.8) | 6.2 (6.0 to 6.4) | 13.2 (13.0 to 13.4) |

| | Asian | Black | Latino/Hispanic | White |
|---|---|---|---|---|
| Hypertension | 21.0 (20.3 to 21.7) | 35.0 (34.5 to 35.6) | 20.0 (19.6 to 20.4) | 28.9 (28.7 to 29.2) |
| Kidney disease | 1.1 (1.0 to 1.3) | 2.2 (2.1 to 2.4) | 1.8 (1.7 to 1.9) | 1.7 (1.7 to 1.8) |
| Liver disease | 1.4 (1.2 to 1.6) | 1.1 (1.0 to 1.2) | 1.7 (1.6 to 1.8) | 1.4 (1.4 to 1.5) |
| Stroke | 1.5 (1.4 to 1.7) | 3.4 (3.3 to 3.6) | 1.7 (1.6 to 1.8) | 2.8 (2.7 to 2.8) |
| Number of concurrent conditions | | | | |
| 0 | 66.9 (66.1 to 67.8) | 51.0 (50.4 to 51.5) | 65.7 (65.3 to 66.2) | 51.3 (51.0 to 51.5) |
| 1 | 21.4 (20.7 to 22.1) | 27.6 (27.2 to 28.0) | 12.1 (20.8 to 21.5) | 27.3 (27.1 to 27.4) |
| ≥2 | 11.7 (11.2 to 12.3) | 21.4 (21.0 to 21.9) | 13.2 (12.8 to 13.5) | 21.5 (21.3 to 21.7) |

Data are presented as % (95% CI) for categorical variables and median (P25-P75) for continuous variables. All percentages are weighted and unadjusted.

[a] The annual family income was categorized into low income and middle/high income relative to the respective year's

federal poverty level from the US Census Bureau (<200% and ≥200%, respectively). The weighted proportion of individuals with low income was estimated using multiple imputation.

[b] Based on the US Census Bureau recognized region of the housing unit where the survey participant was interviewed.

Abbreviations: BMI, body mass index; CI, confidence interval; COPD, chronic obstructive pulmonary disease; GED, general equivalency diploma.

**S1 Figure. Study Population Flowchart.**

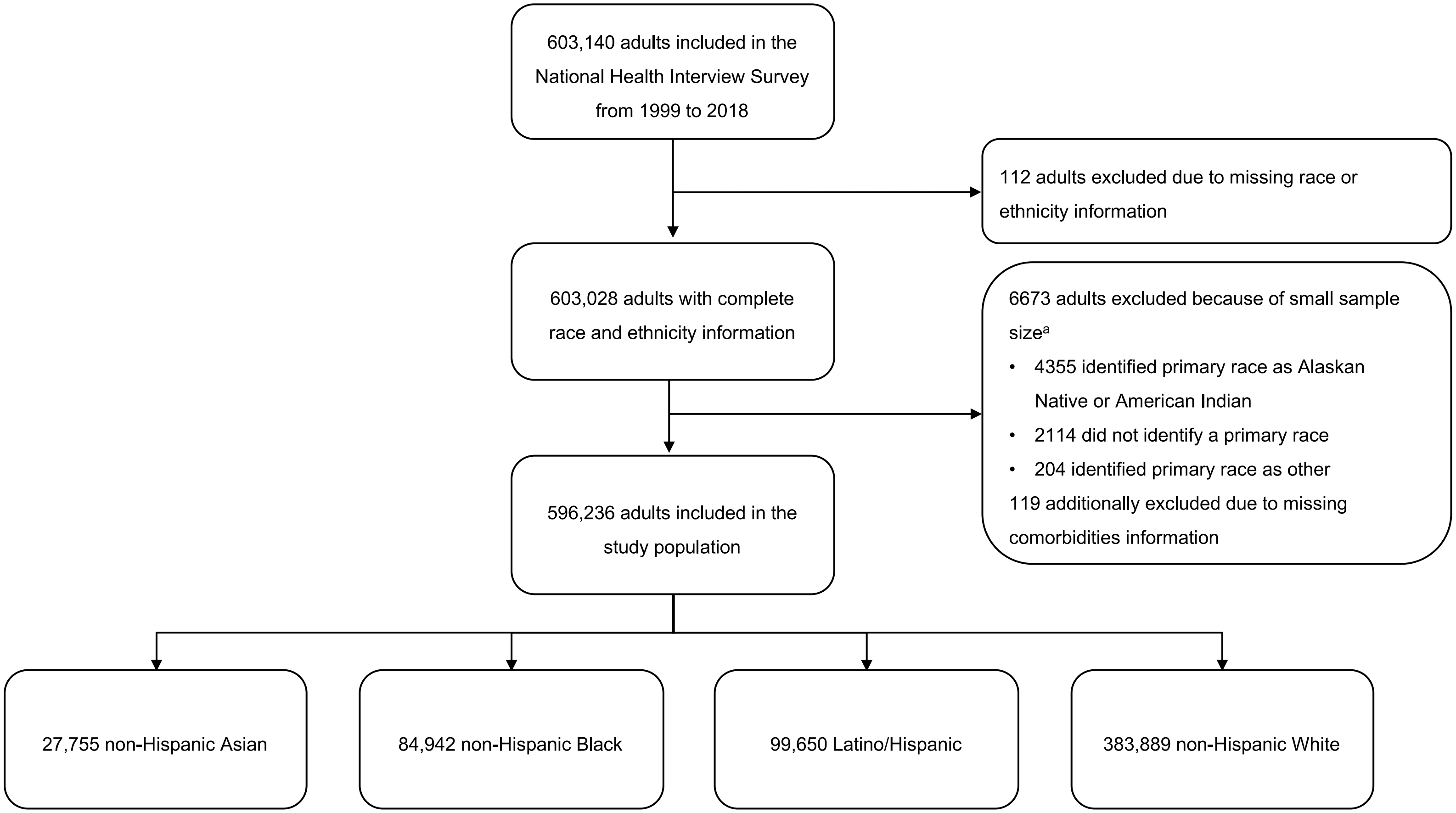


The mutually exclusive racial/ethnic subgroups were created based on the self-reported primary race and ethnicity combination.
[a] These excluded individuals also did not identify as Latino/Hispanic.

**S2 Figure. Unadjusted Age Distribution by Race and Ethnicity.**

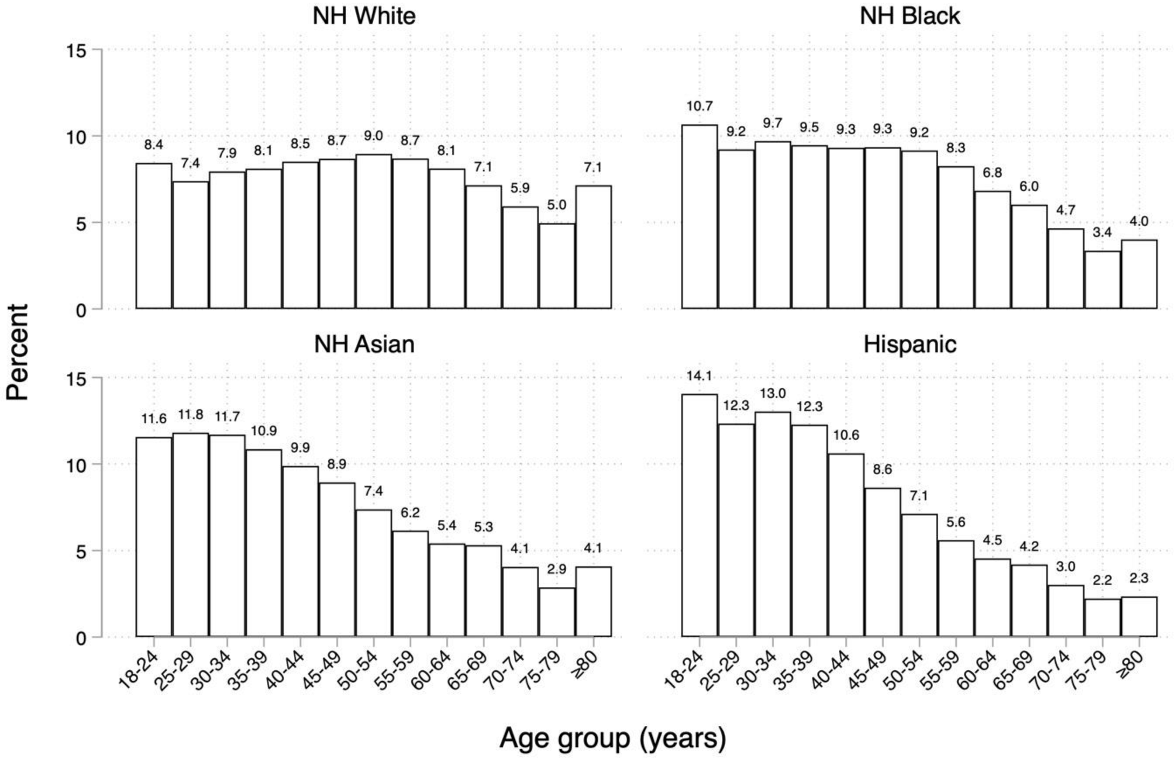


Abbreviations: NH, non-Hispanic.

**S3 Figure. Unadjusted Distribution of Number of Chronic Conditions by Race and Ethnicity**

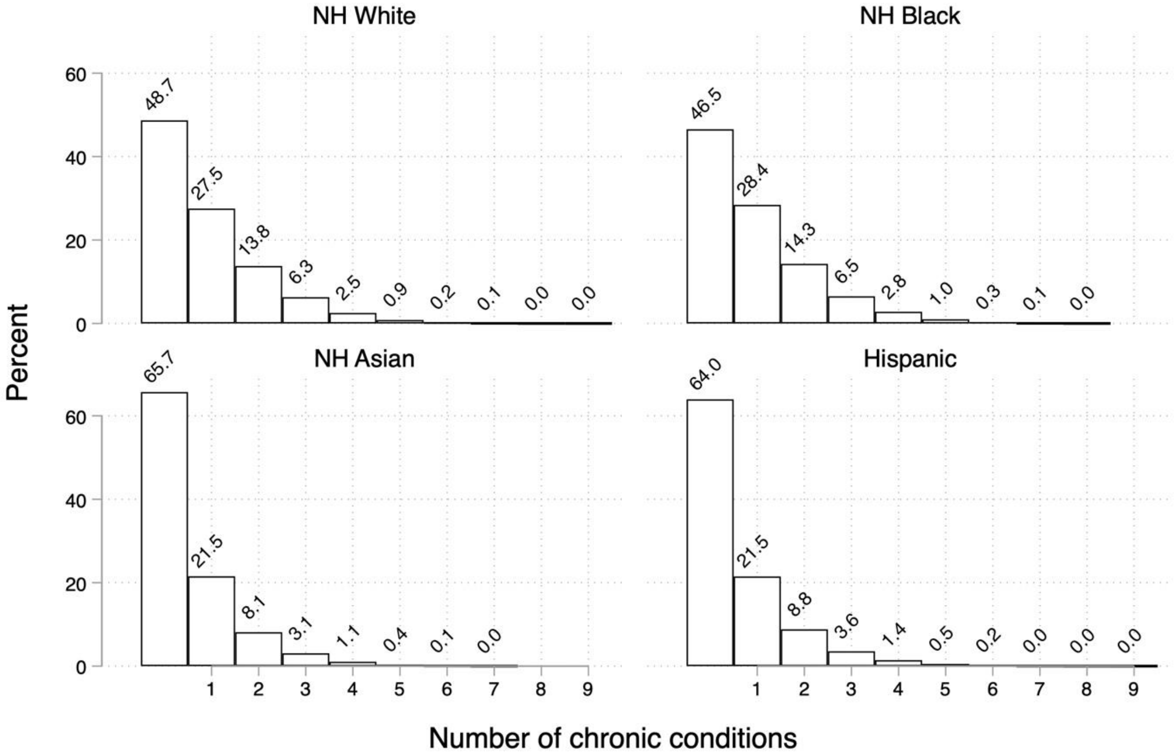

**S4 Figure. Trends in Adjusted Prevalence of Individual Conditions by Race and Ethnicity, 1999-2018.**

**A) Asthma**

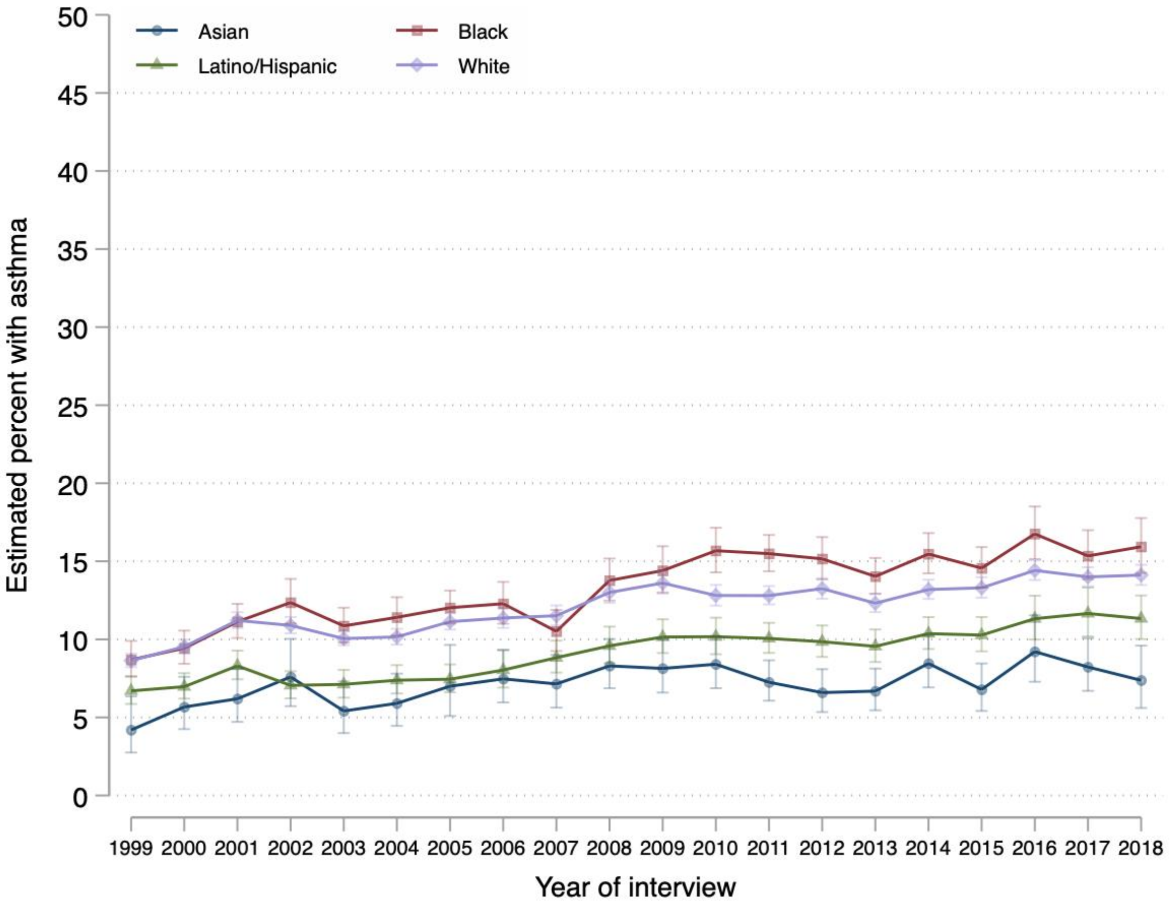

**B) Cancer**

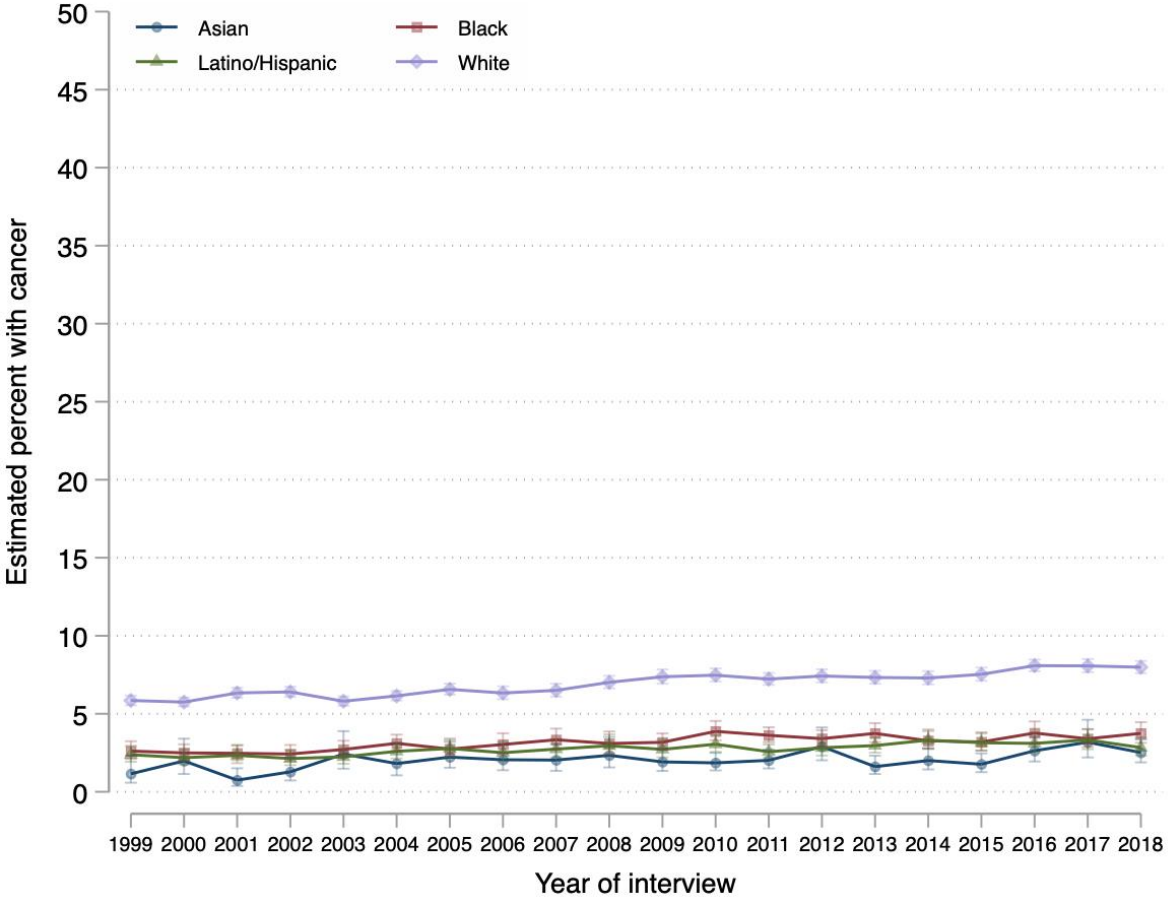

Asian
Black
Latino/Hispanic
White
Estimated percent with cancer
50
45
40
35
30
25
20
15
10
5
0
1999 2000 2001 2002 2003 2004 2005 2006 2007 2008 2009 2010 2011 2012 2013 2014 2015 2016 2017 2018
Year of interview

**C) Chronic kidney disease**

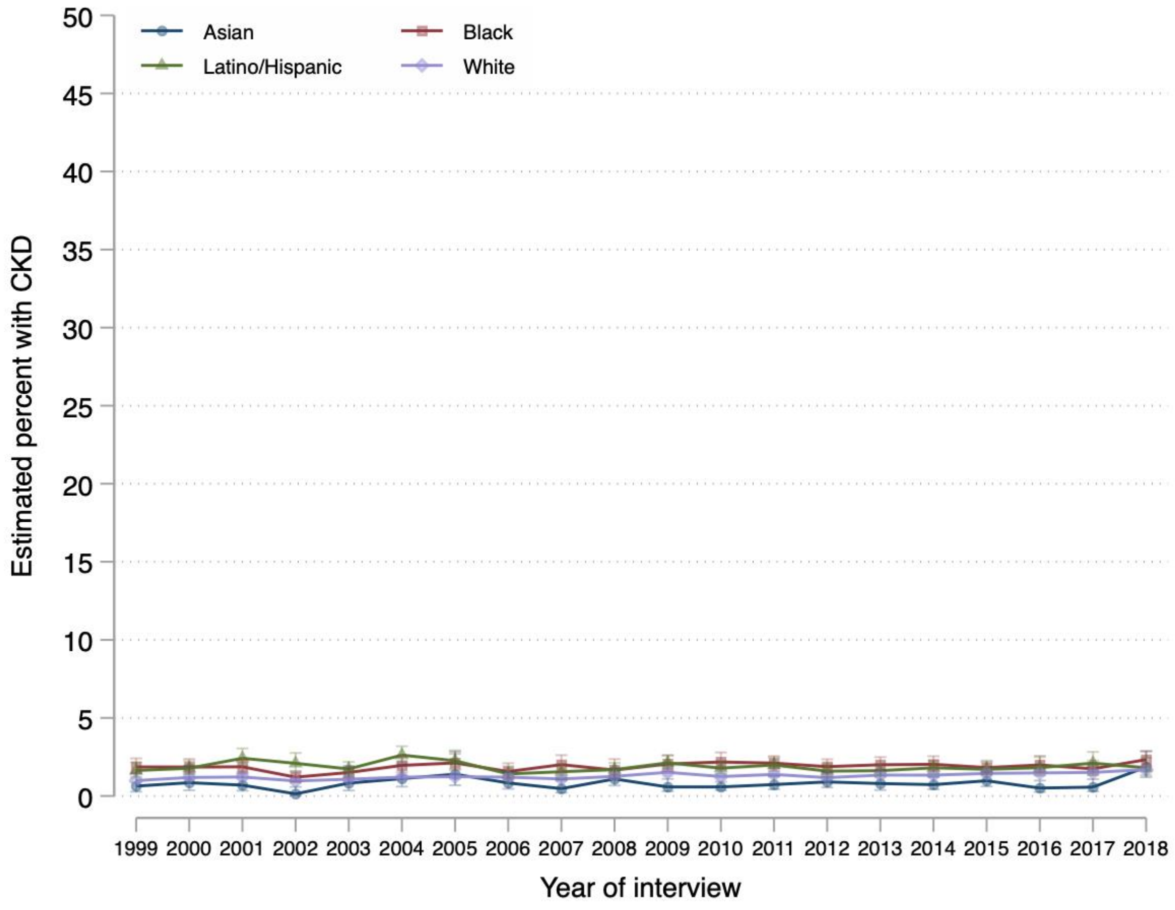

**D) Chronic bronchitis or emphysema**

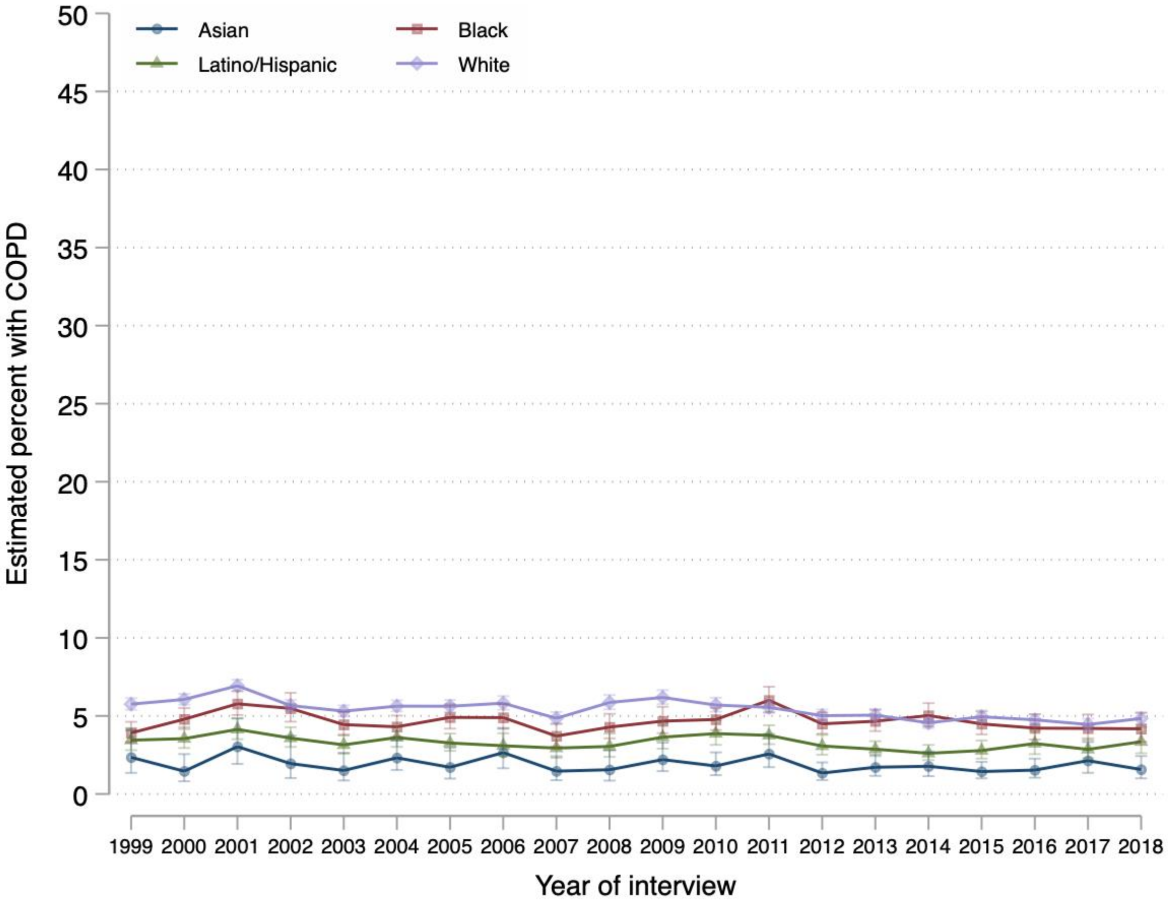

**E) Diabetes**

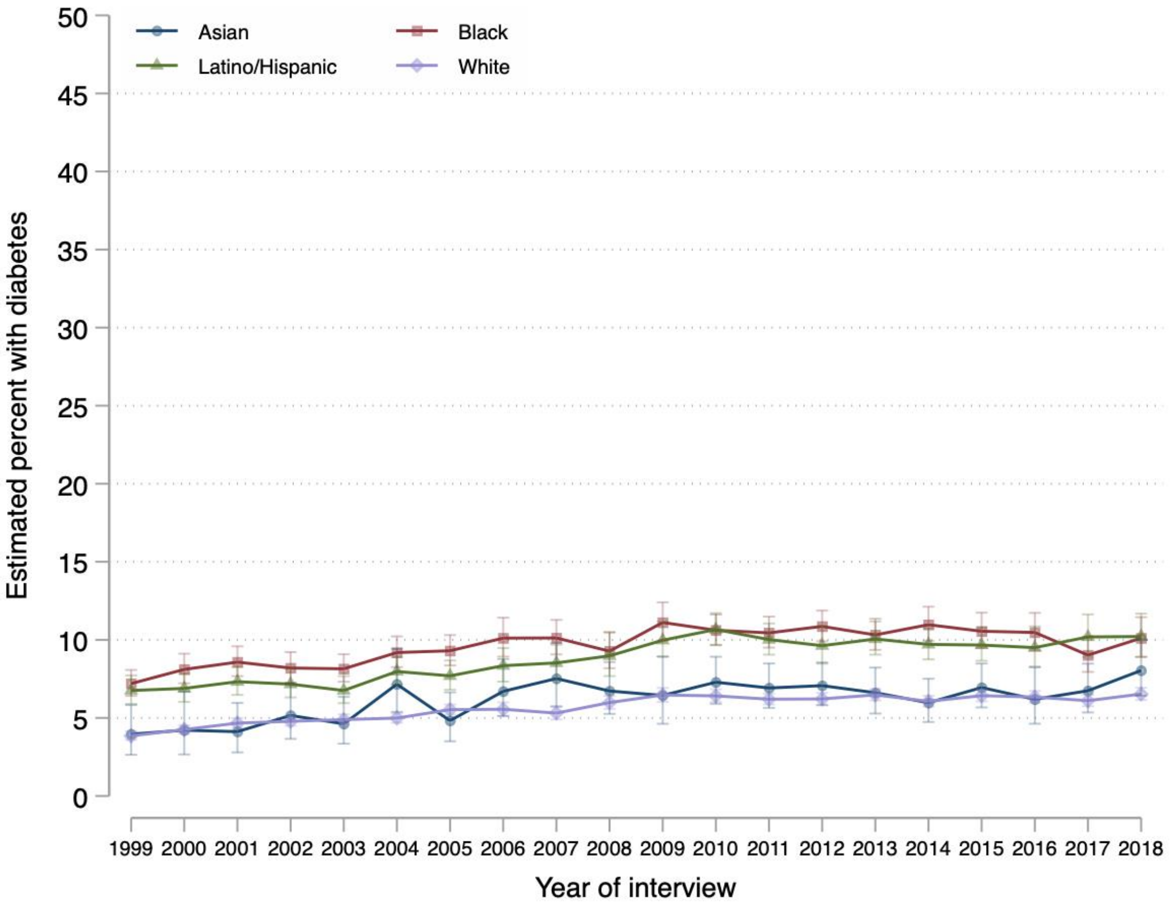
Asian
Black
Latino/Hispanic
White
Estimated percent with diabetes
0
5
10
15
20
25
30
35
40
45
50
1999 2000 2001 2002 2003 2004 2005 2006 2007 2008 2009 2010 2011 2012 2013 2014 2015 2016 2017 2018
Year of interview

**F) Heart disease**

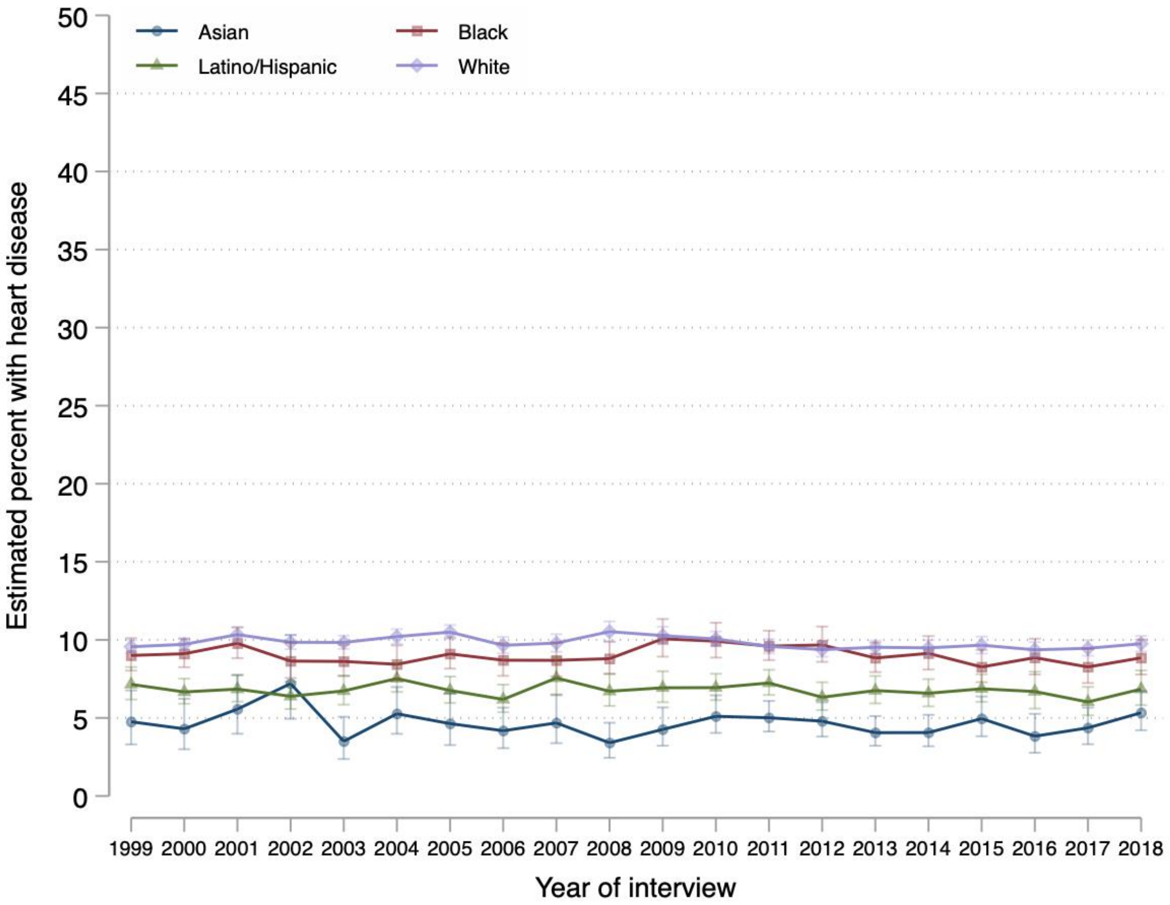

**G) Hypertension**

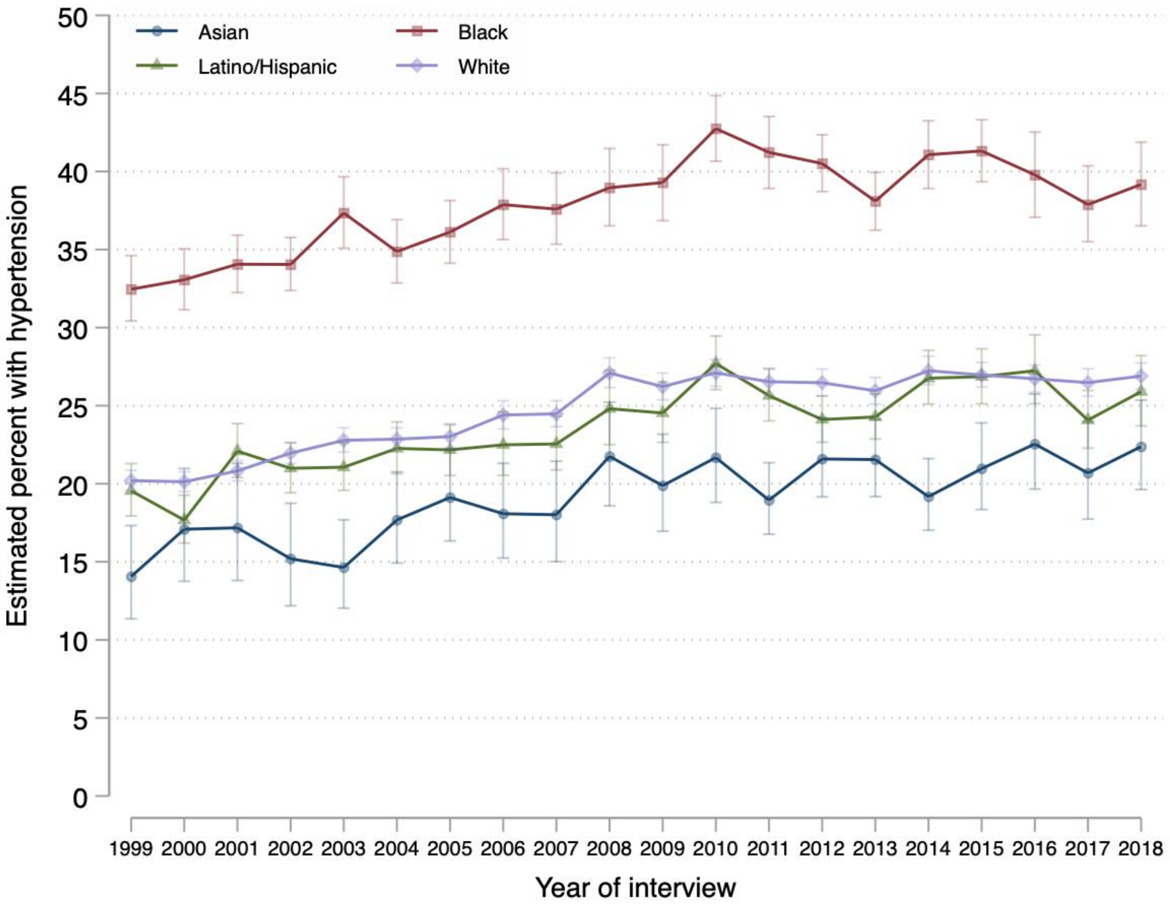

**H) Liver disease**

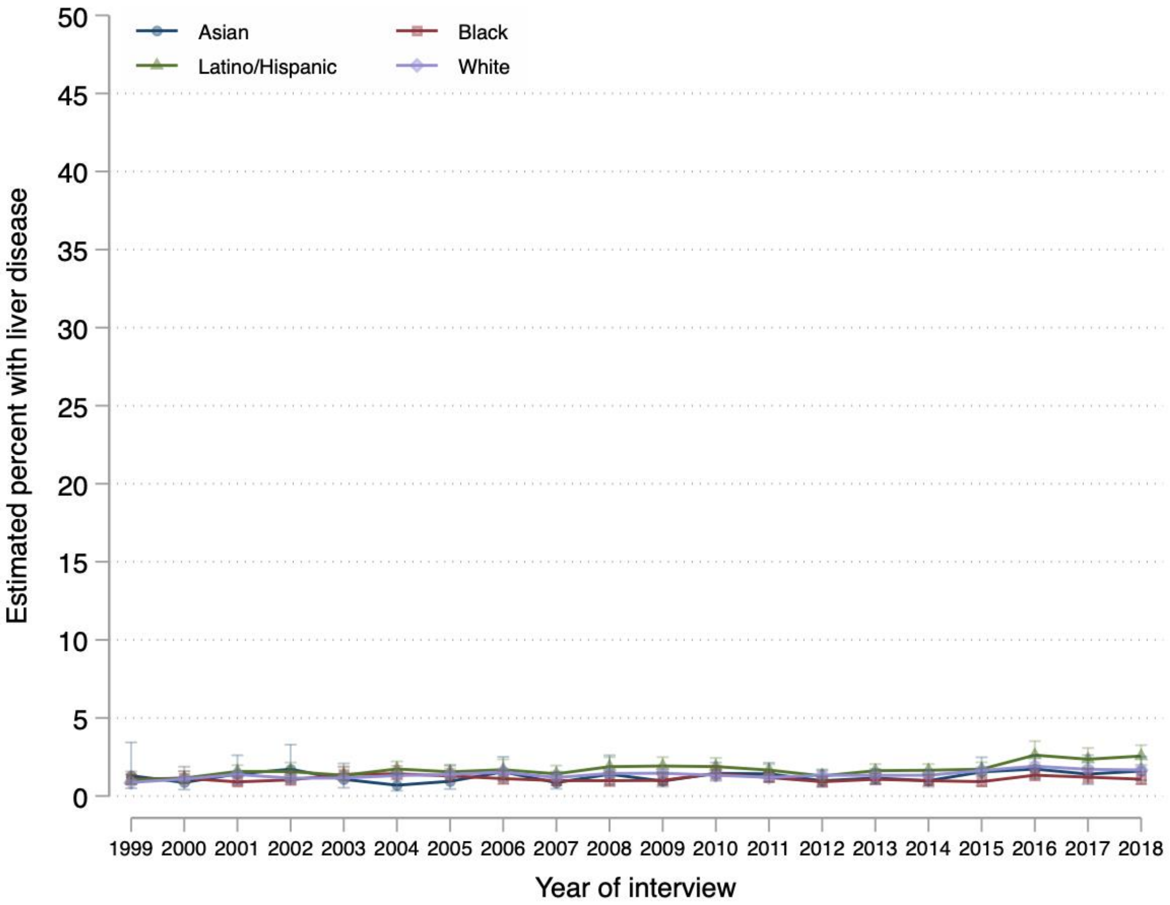

**I) Stroke**

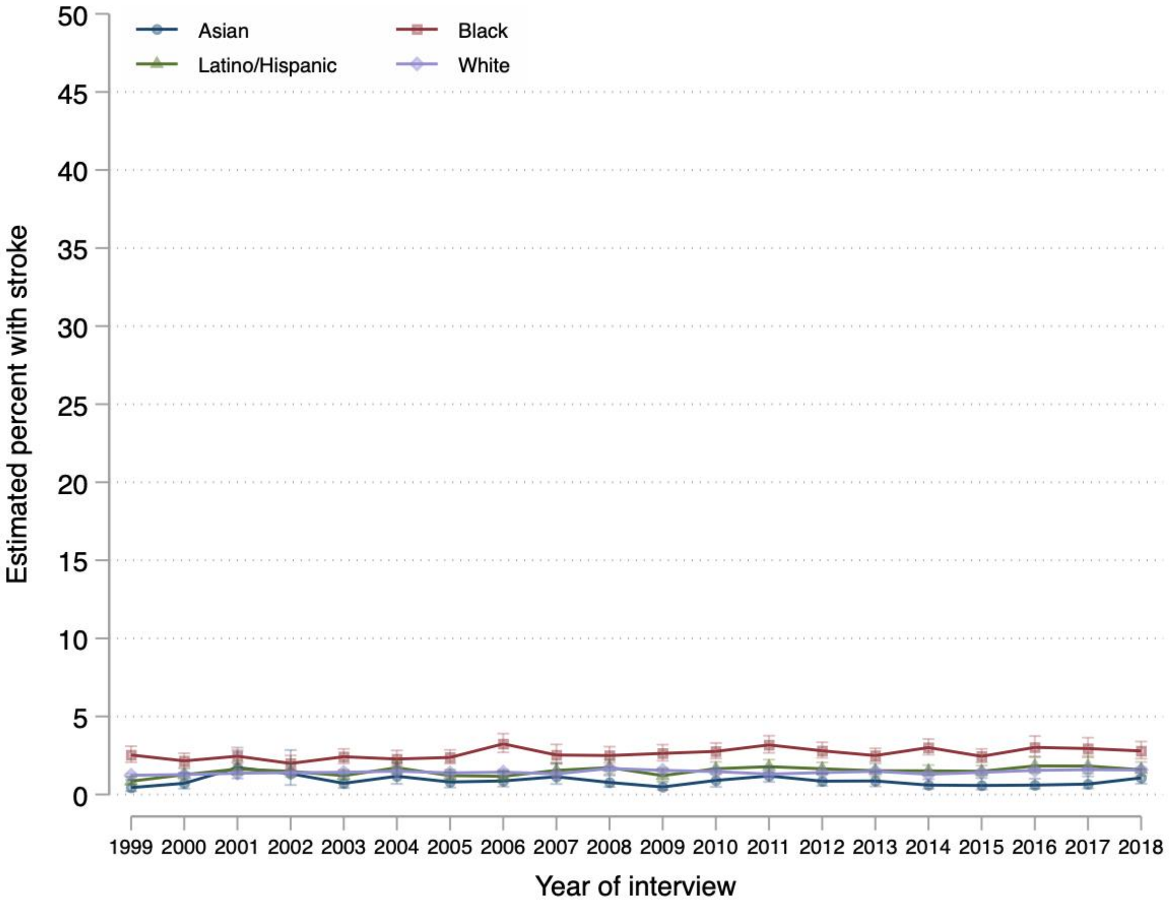


Abbreviations: CKD, chronic kidney disease; COPD, chronic obstructive pulmonary disease

**S5 Figure. Trends in Adjusted Multimorbidity Prevalence by Race, Ethnicity, and Income (A) and by Income Alone (B), 1999-2018.**

**A)**

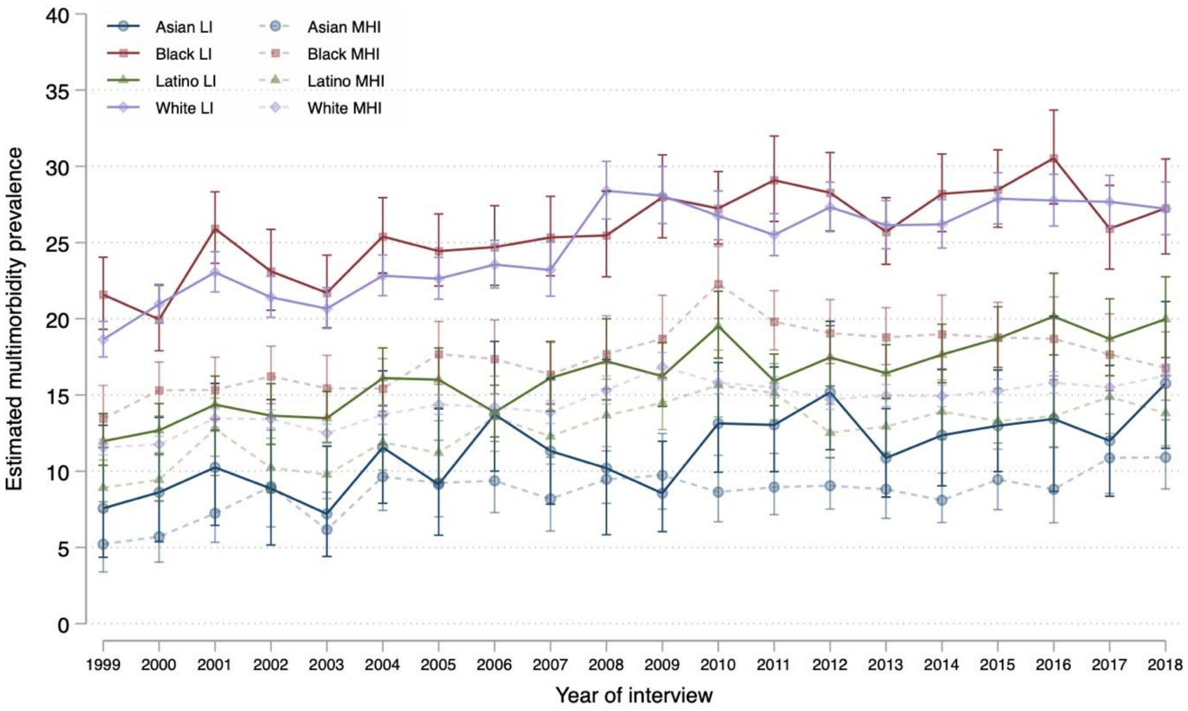

**B)**

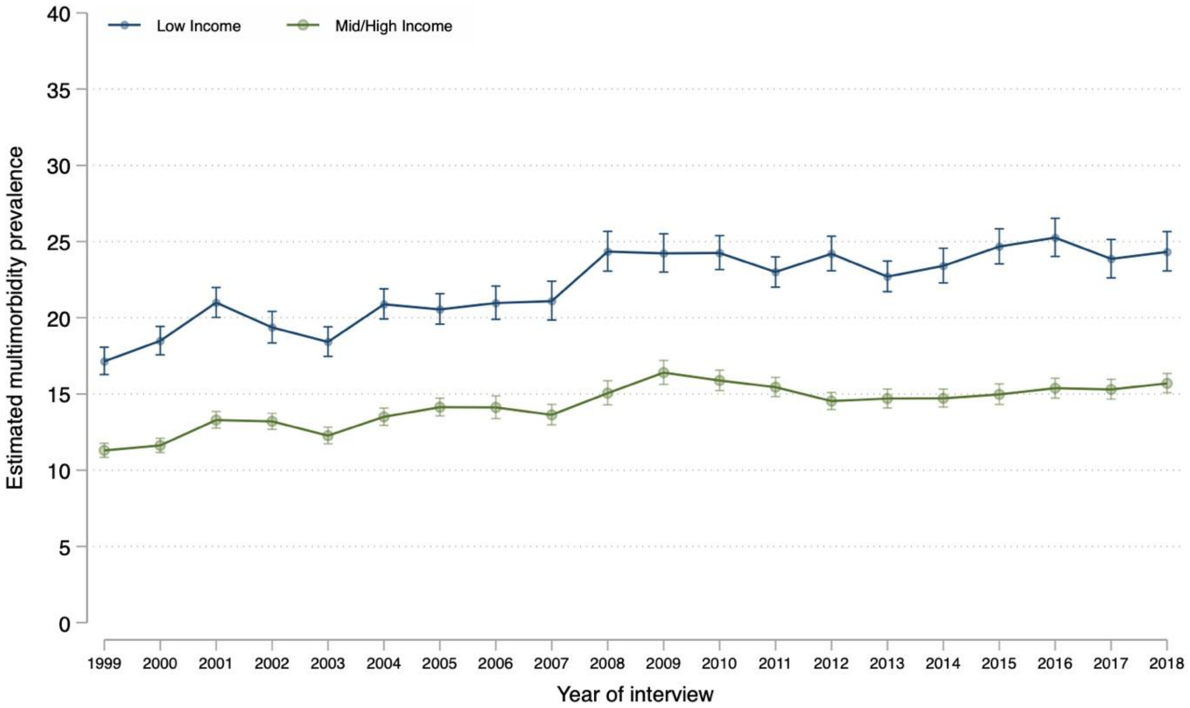


Legend: Annual family income was categorized relative to the respective year's Federal Poverty Limit from the US Census Bureau into middle/high income (≥200%) and low income (<200%). The weighted proportion of individuals with annual income <200% of the Federal Poverty Limit was estimated using multiple imputation.

Abbreviations: LI, low income; MHI, middle/high income.

**S6 Figure.** Number of Chronic Conditions by Age Among Asian, Black, Latino/Hispanic, and White People.

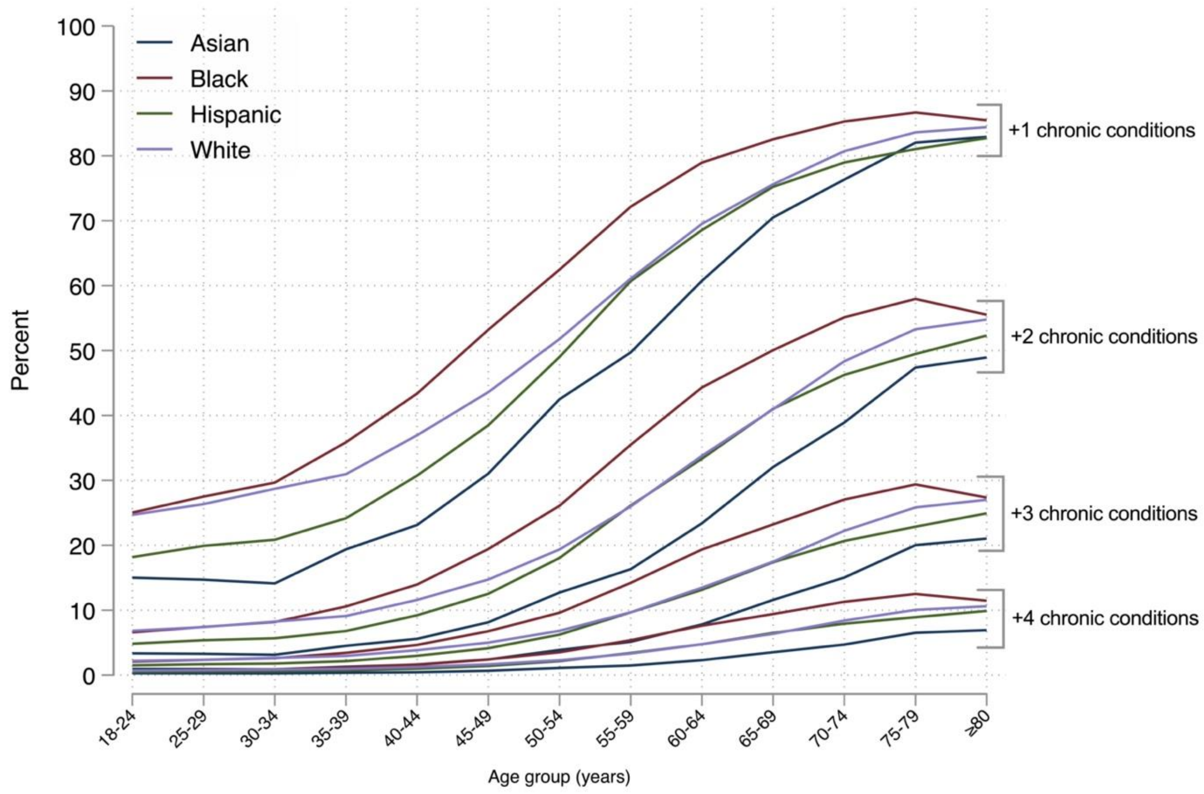


Legend: Data source is the National Health Interview Survey from 1999 to 2018.

Estimates were obtained using ordered logistic regression models, adjusting for sex and region (details in Methods).

**S7 Figure. Adjusted Multimorbidity Prevalence by Age, Race, Ethnicity, and Income (A), and by Age and Income (B).**

**A)**

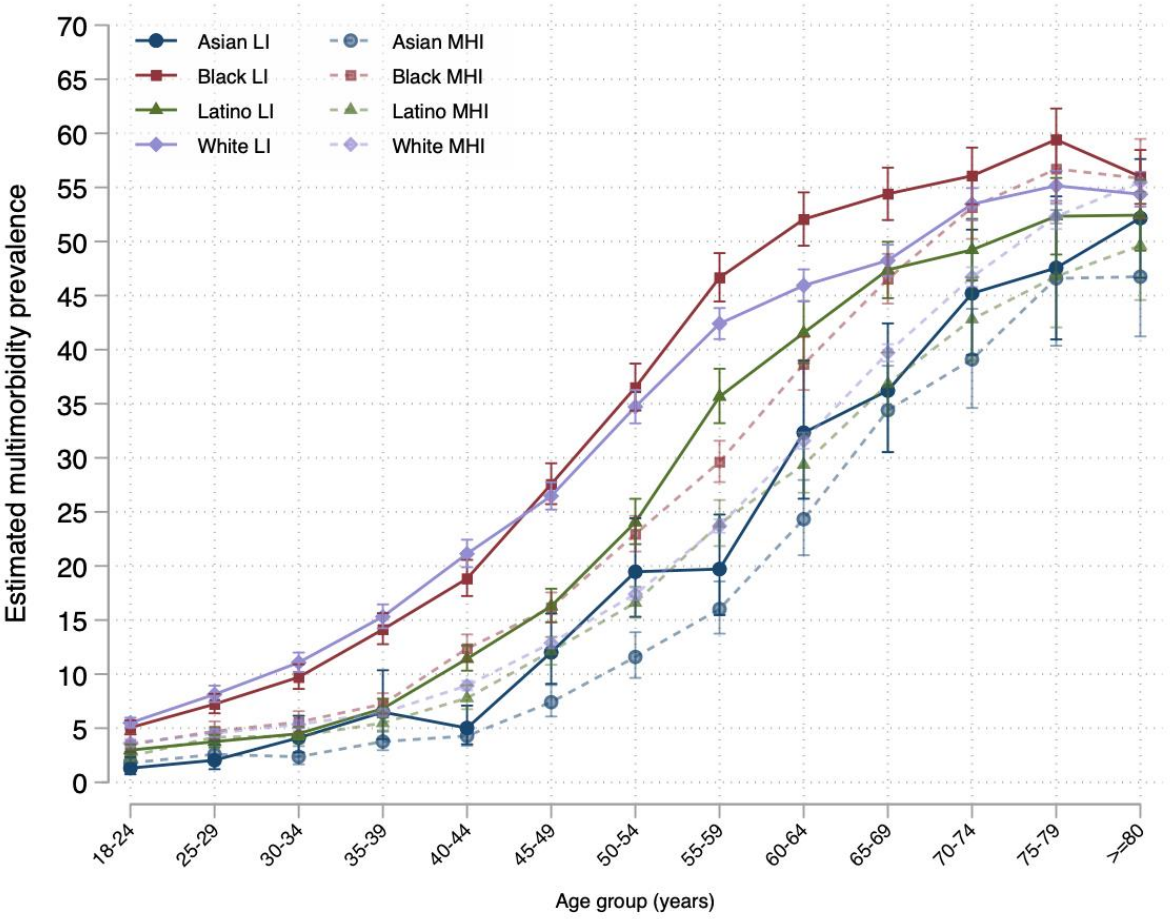

**B)**

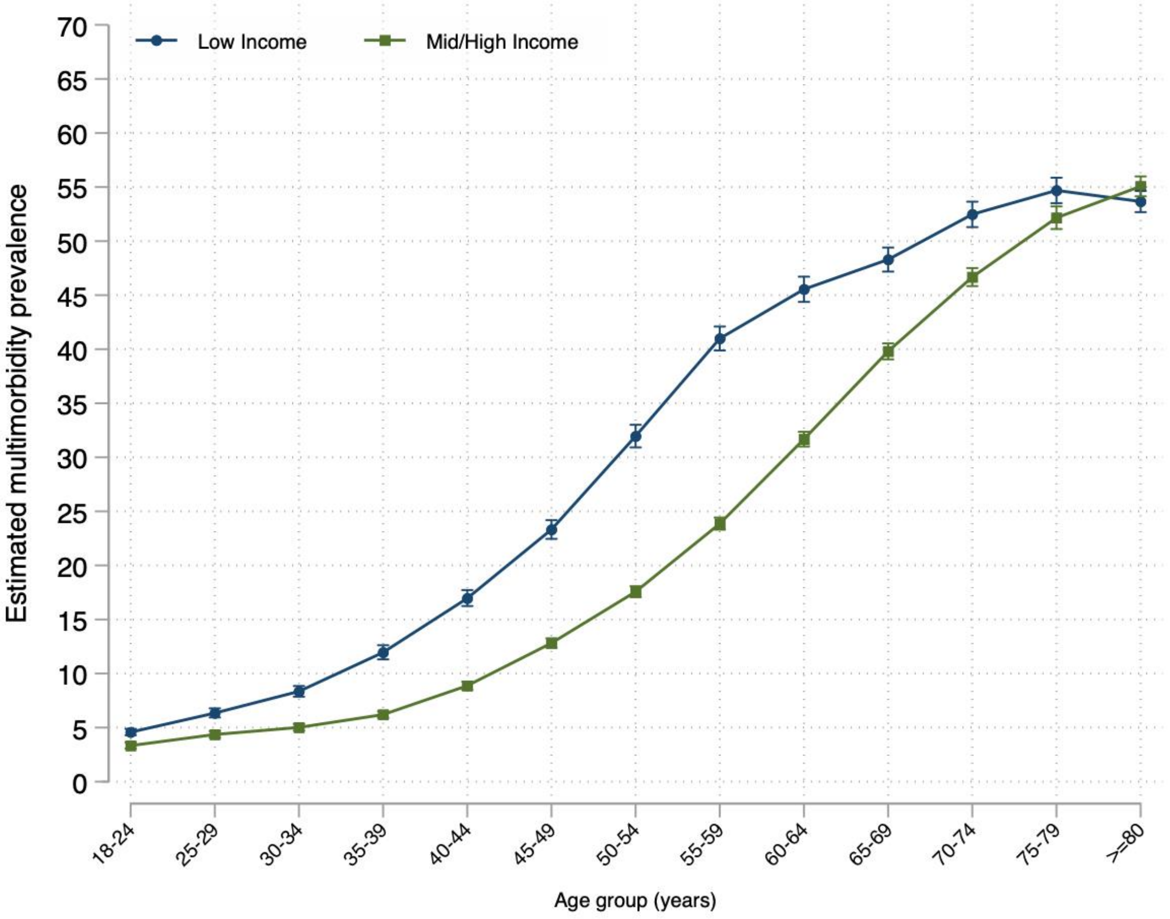


Annual family income was categorized relative to the respective year's Federal Poverty Limit from the US Census Bureau into middle/high income (≥200%) and low income (<200%). The weighted proportion of individuals with annual income <200% of the Federal Poverty Limit was estimated using multiple imputation.

Abbreviations: LI, low income; MHI, middle/high income.

**S8 Figure. Sensitivity Analysis Including Arthritis: Trends in Adjusted Multimorbidity Prevalence by Race and Ethnicity, 2002-2018.**

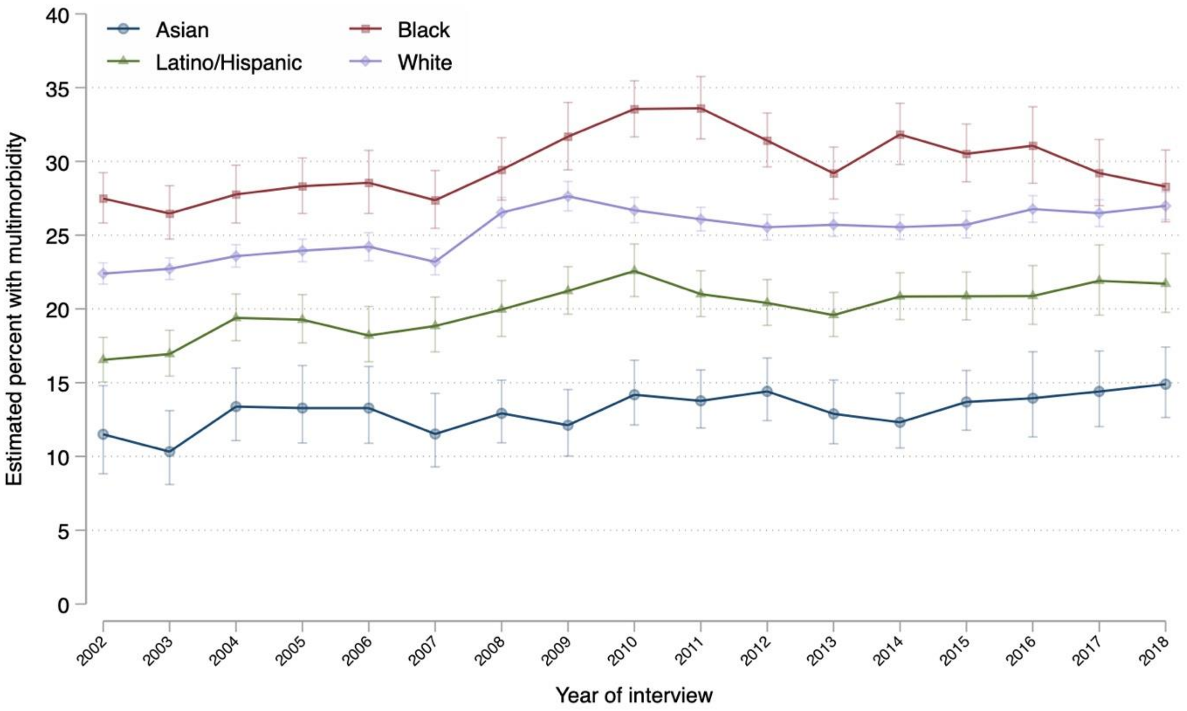


| | Asian | Black | Latino/Hispanic | White |
|---|---|---|---|---|
| **Multimorbidity** | Percentage points (95% CI), p value | Percentage points (95% CI), p value | Percentage points (95% CI), p value | Percentage points (95% CI), p value |
| **Annualized rate of change in prevalence** | +0.15 (+0.04 to +0.26), 0.01 | +0.24 (+0.03 to +0.44), 0.03 | +0.26 (+0.15 to +0.38), <0.001 | +0.26 (+0.16 to +0.36), <0.001 |
| **Absolute change in prevalence, 2002–2018** | +3.40 (-0.43 to +7.22), 0.08 | +0.80 (-2.17 to +3.78), 0.60 | +5.16 (+2.66 to +7.66), <0.001 | +4.59 (+3.42 to +5.76), <0.001 |
| **Difference with White, 2002** | -10.90 (-13.97 to -7.82), <0.001 | +5.09 (+3.23 to +6.95), <0.001 | -5.85 (-7.53 to -4.17), <0.001 | - |
| **Difference with White, 2018** | -12.09 (-14.65 to -9.53), <0.001 | +1.31 (-1.30 to +3.91), 0.33 | -5.28 (-7.47 to -3.09), <0.001 | - |
| **Absolute change in difference with White, 2002–2018** | -1.19 (-5.19 to +2.81), 0.56 | -3.78 (-6.98 to -0.58), 0.02 | +0.57 (-2.19 to +3.33), 0.68 | - |

Legend: Data source is the National Health Interview Survey from years 2002 to 2018. For change in prevalence and change in difference: a positive sign (+) means the prevalence of multimorbidity (or its difference with White people) increased and a negative sign (-) means it decreased. Multimorbidity prevalence was adjusted by age, sex, and region.
Abbreviations: CI, confidence interval

**S9 Figure. Sensitivity Analysis Including Arthritis: Number of Chronic Conditions by Age Among Asian, Black, Latino/Hispanic, and White People.**

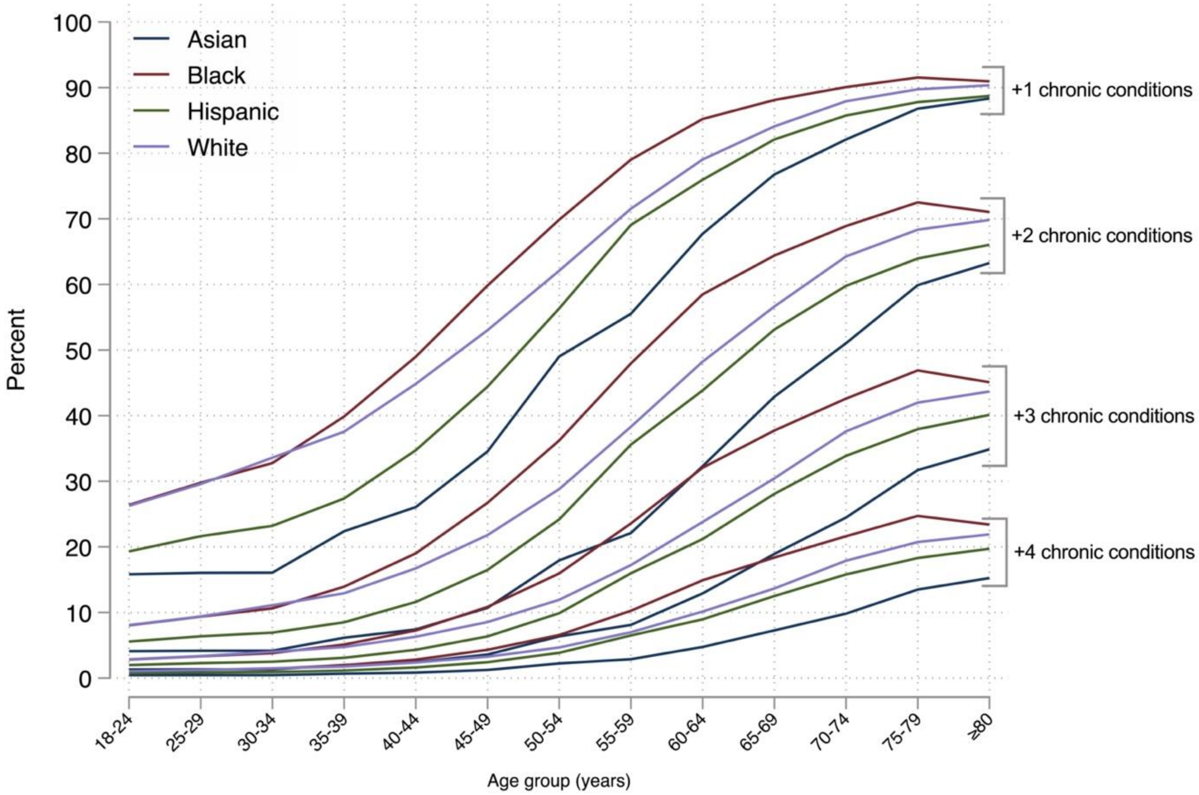

**S10 Figure. Sensitivity Analysis Including Arthritis: Adjusted Multimorbidity Prevalence by Age, Race, and Ethnicity (A), and Prevalence Differences by Age, Race, and Ethnicity (B).**

**A)**

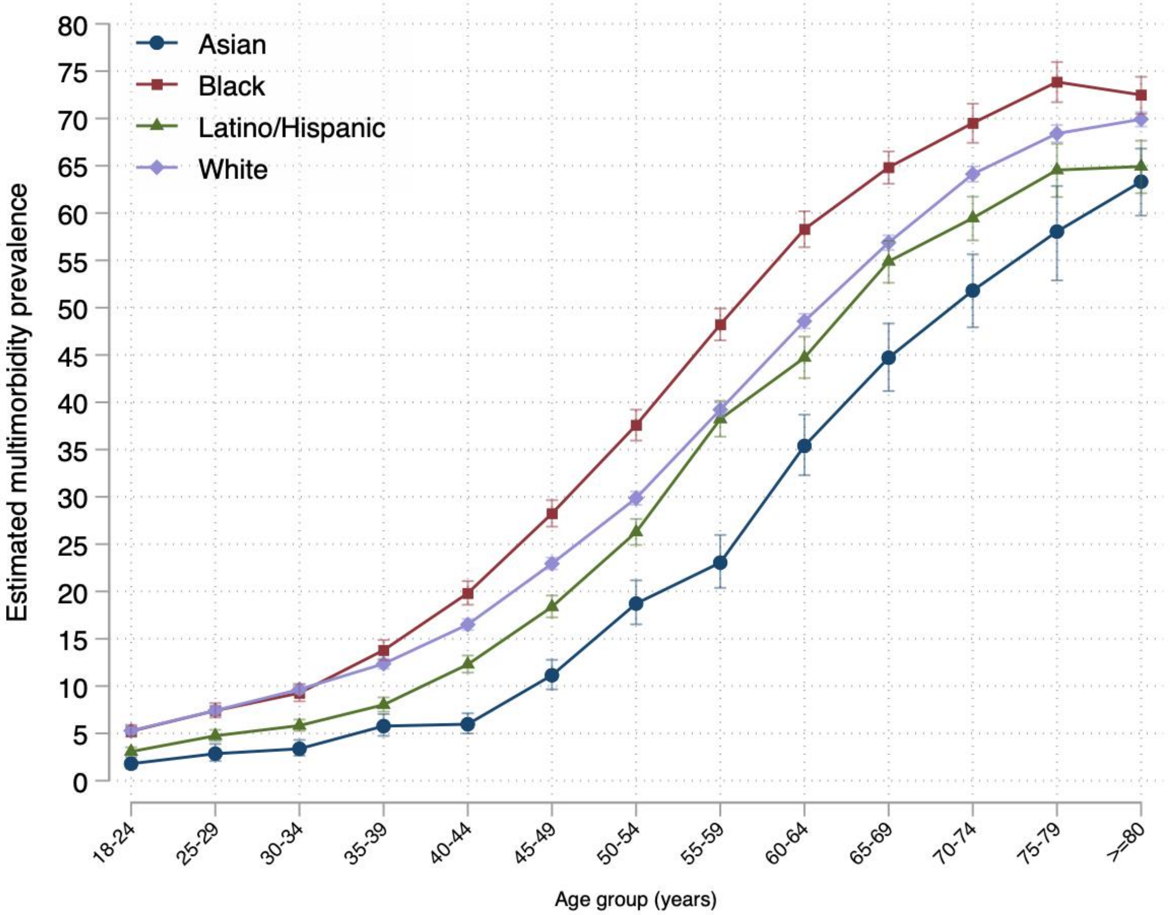

**B)**

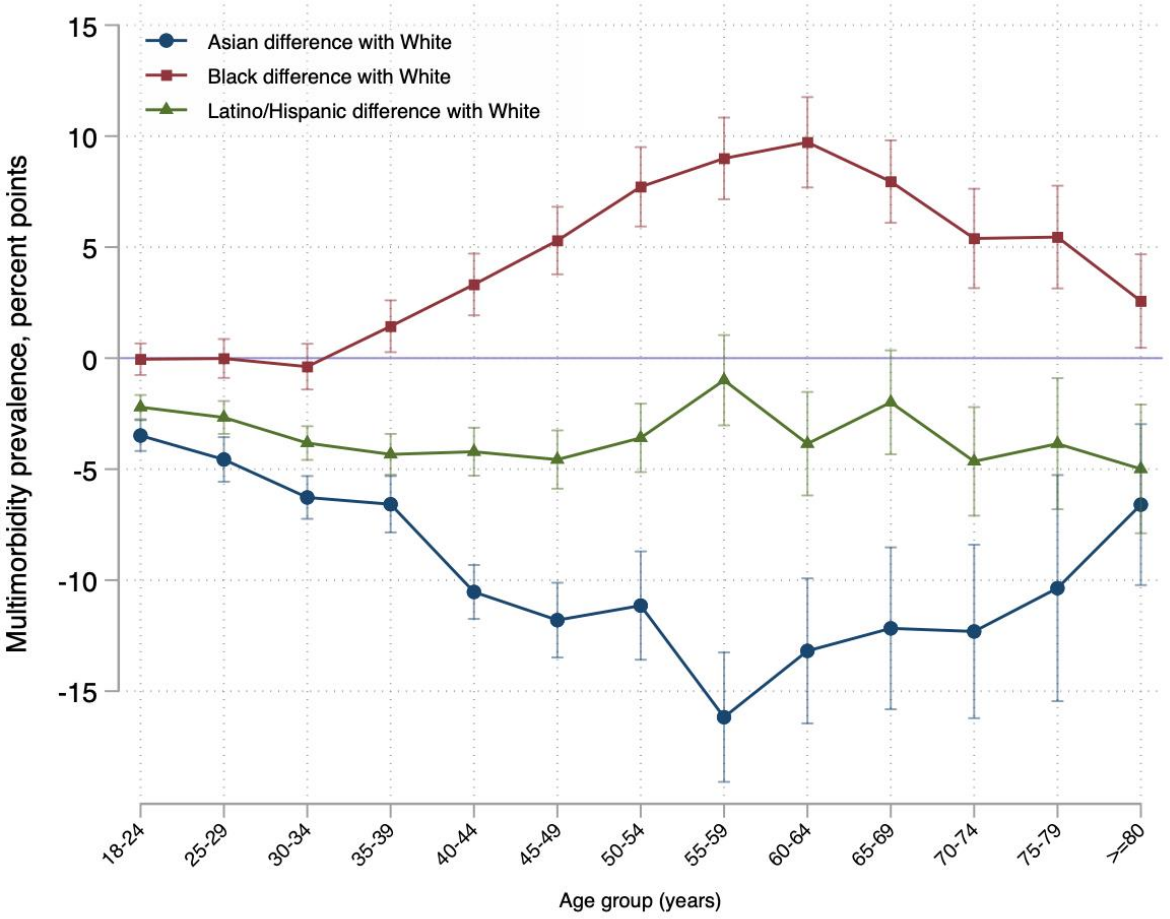
Asian difference with White
Black difference with White
Latino/Hispanic difference with White
Multimorbidity prevalence, percent points
15
10
5
0
-5
-10
-15
18-24
25-29
30-34
35-39
40-44
45-49
50-54
55-59
60-64
65-69
70-74
75-79
>=80
Age group (years)

**S11 Figure. Sensitivity Analysis: Trends in Adjusted Prevalence of Arthritis by Race and Ethnicity, 2002-2018.**

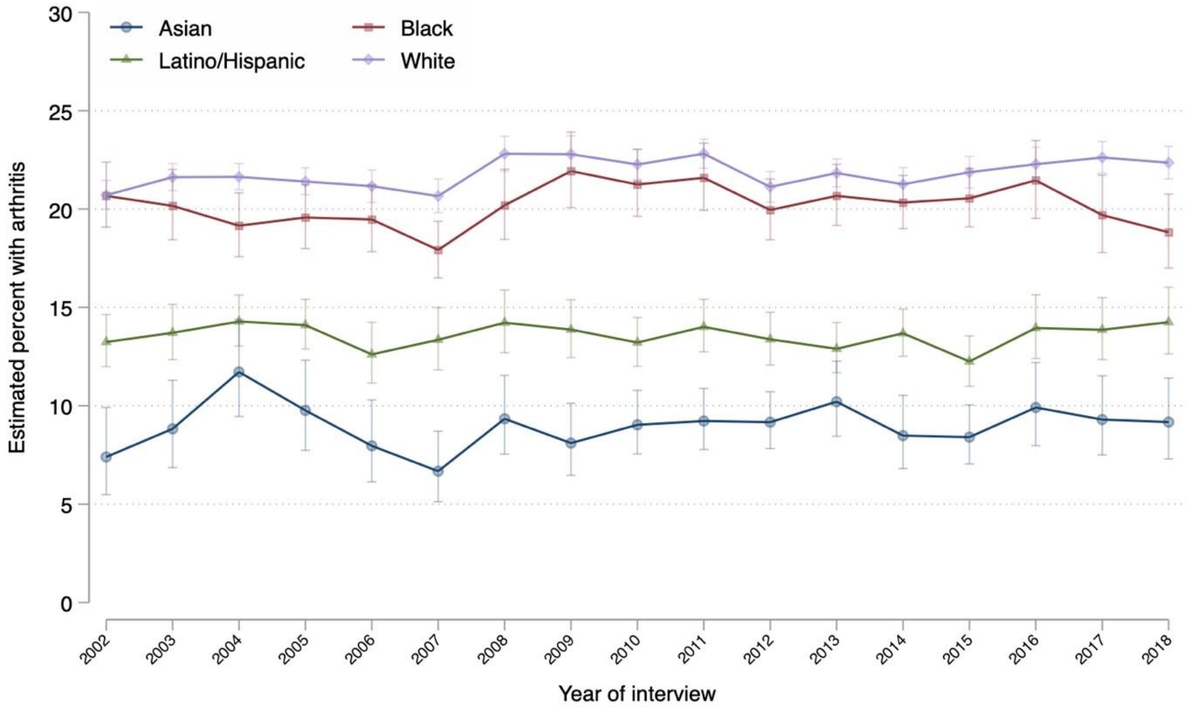
